# Pathogenic variants in the cohesin loader subunit MAU2 lead to a new Cornelia de Lange Syndrome subtype

**DOI:** 10.1101/2025.11.16.25339042

**Authors:** Ilaria Parenti, Alina Hesters, Marta Gil-Salvador, Laura Duffy, Deniz Kanber, Jasmin Beygo, Jennifer Kerkhof, Laura Steenpaß, Elsa Leitão, Julia Woestefeld, Philip M Boone, Emeline M Kao, Lama Alabdi, Hesham M Aldhalaan, Fowzan S Alkuraya, Muneera J Alshammari, Stylianos E Antonarakis, Donald Basel, Kevin Cassinari, Laurana de Polli Cellin, Amanda R Clause, Alexander Augusto de Lima Jorge, Andréa de Castro Leal, Stephan C Collins, Benjamin Durand, Juliane Eckhold, Mais O Hashem, Parul Jayakar, Arif O Khan, Kohji Kato, Regina Kubica, Gholson J Lyon, Elaine Marchi, Julie McCarrier, Lara K Kimmig, Seiji Mizuno, Gael Nicolas, Yosuke Nishio, Tomoo Ogi, Juan Pié, Jordyn Prell, Beatriz Puisac, Feliciano J Ramos, Emmanuelle Ranza, Claire Redin, Eric Rush, Shinji Saitoh, Hanan E Shamseldin, Susan Starling, Esteban Astiazaran-Symonds, Sara Taher, Alma Kuechler, Bekim Sadikovic, Binnaz Yalcin, Kerstin S Wendt, Frank J Kaiser

## Abstract

The role of the cohesin complex depends on the cohesin loader proteins NIPBL and MAU2. While *NIPBL* variants are a major cause of Cornelia de Lange Syndrome (CdLS), the role of *MAU2* in disease is unclear. We describe 18 individuals carrying 15 heterozygous *MAU2* variants and demonstrate pathogenicity through functional analyses. MAU2 in-frame variants predominantly impair NIPBL–MAU2 interaction, whereas truncating variants cause *MAU2* haploinsufficiency and lead to NIPBL reduction. Most patients exhibit a DNA methylation profile compatible with the CdLS episignature. We also identified two *MAU2*-specific episignatures that reflect variant-dependent molecular consequences. Affected individuals display a wide range of phenotypes, from classic CdLS to milder presentations, with short stature and microcephaly as consistent features. A heterozygous *Mau2* knockout mouse model recapitulated these traits, confirming the causal role of *MAU2* disruption *in vivo.* Our study establishes *MAU2* as a new CdLS-associated gene and delineates a *MAU2*-related chromatinopathy with variable expressivity.

## Introduction

Cornelia de Lange Syndrome (CdLS, OMIM #122470, #300590, #610759, #614701, #300882, and #620568) is a rare multisystem neurodevelopmental disorder with an estimated incidence between 1:10,000 and 1:30,000 live births^1^. Craniofacial features are a hallmark of the syndrome and comprise synophrys, thick and arched eyebrows, a depressed nasal bridge with anteverted nares, a long and smooth philtrum, and a thin upper lip with downturned corners^1^. Additional common signs include developmental delay, intellectual disability, behavioral anomalies, growth retardation, microcephaly, limb anomalies, and hirsutism. The number and severity of these clinical features vary among individuals, and additional organ or system involvement may occur, underscoring the broad clinical variability characteristic of CdLS^1^. An international consensus statement has developed a scoring system designed to standardize the clinical assessment of the syndrome, thereby facilitating the diagnosis and the evaluation of disease severity^1^.

CdLS is primarily associated with pathogenic variants in subunits or regulators of cohesin, an evolutionarily conserved multisubunit complex that plays key roles in sister chromatid cohesion, DNA repair, chromatin architecture, and transcriptional regulation^2^. It is primarily the disruption of the latter function that appears to underlie the development of CdLS. Accordingly, cells from patients with CdLS do not exhibit cohesion defects but rather show global dysregulation of chromatin conformation and transcription. This dysregulation provides the basis for classifying CdLS as a chromatinopathy^3, 4^. Pathogenic variants in six different genes are most frequently reported in CdLS patients^1^. Among these, the cohesin loader subunit NIPBL (OMIM #608667) is the primary contributor, with disease-causing variants identified in approximately 70% of affected individuals. Pathogenic variants in this gene are often associated with classic manifestations of the disease^1^. Up to 15% of cases are attributed to pathogenic variants in the *SMC1A*, *SMC3*, *RAD21*, *HDAC8*, and *BRD4* genes (OMIM #300040, #606062, #606462, #300269, and #608749)^1, 5^. Individuals with pathogenic variants in these genes are characterized by a wide clinical expressivity, including mild presentations as well as non-classical phenotypes^1^. Sporadic reports of CdLS patients with disease-causing variants in non-cohesin proteins with roles in transcriptional regulation and chromatin remodeling further strengthen the classification of CdLS as chromatinopathy^6, 7^.

The primary CdLS-associated protein, NIPBL, forms a heterodimeric complex with MAU2 (OMIM #614560)^8^. MAU2 enwraps the N-terminal region of NIPBL, promoting its proper folding and enabling the assembly of a functional cohesin loader complex^9^. This interaction is crucial for the stability of both proteins and the depletion of either component ultimately results in the downregulation of the other^10^. Despite the close functional interdependence between NIPBL and MAU2, the frequency of pathogenic variants differs strikingly: while 683 *NIPBL* variants are reported in the Human Gene Mutation Database (HGMD^11^), only two have been described in *MAU2*. We have previously reported the first patient with a disease-causing in-frame deletion in *MAU2* and a classic CdLS phenotype^12^. More recently, a second individual carrying a missense substitution was described^13^. This patient presented with a CdLS score warranting molecular testing, but with a rather atypical clinical presentation^13^. In addition, few microdeletions at 19p13.11-p12 have been identified in individuals with neurodevelopmental delay. Some of these deletions encompass *MAU2* along with additional genes, making it challenging to delineate the specific contribution of *MAU2* to the resulting phenotypes^14^. In view of the available data, the precise role of *MAU2* in the context of disease onset remained unclear. In this study, we report 18 patients with 15 heterozygous variants in *MAU2*. Through a combination of detailed phenotyping, episignature analysis, functional assays, and the generation of a mouse model, we characterize this cohort and assess the molecular and phenotypic consequences of *MAU2* variants. Our findings demonstrate that the majority of these variants give rise to a new CdLS subtype mainly characterized by short stature and microcephaly. Based on these observations, we propose that *MAU2* should be recognized as a new CdLS-associated gene.

## Results

### Short-reads high-throughput sequencing identifies novel *MAU2* variants

We report 15 distinct heterozygous variants in the *MAU2* gene across 18 patients. Based on gnomAD v4.1.0^15^ constraint metrics, *MAU2* is predicted to be intolerant to both loss-of-function and missense variation, with a pLI score of 1 and a Z-score of 4.86. Reflecting this constraint, we identified both truncating and in-frame variants in our cohort. The collected variants comprise five missense substitutions (p.(Cys50Ser), p.(Leu381Pro), p.(Cys394Tyr), p.(Leu528Phe), and p.(Asp534His)), four in-frame deletions (p.(Val54_Pro56del), p.(Ala309_Lys322del), p.(Lys322del), and p.(Gln310_Ala316del)), one in-frame delins (p.(Ala378_Gln380delinsAsp)), one nonsense variant (p.(Gln538*)), two frameshift duplications (p.(Cys145Leufs*14) and p.(Asp242Glyfs*28)), and two frameshift deletions, one resulting in a premature stop codon (p.(Gln135Argfs*32)), and one resulting in protein elongation (p.(Ala601Leufs*124)) (Figure 1). Only the p.(Cys50Ser) was found in gnomAD v4.1.0 (6 alleles), indicating that the *MAU2* variants reported here are extremely rare in population databases. Notably, the patient harboring the p.(Gln310_Ala316del) in-frame deletion had been previously described by our group^12^. To enable a more comprehensive clinical and molecular characterization of the consequences of the *MAU2* variants, we have also included this individual in the present cohort, where he is designated as Patient 1.

**Figure 1.**
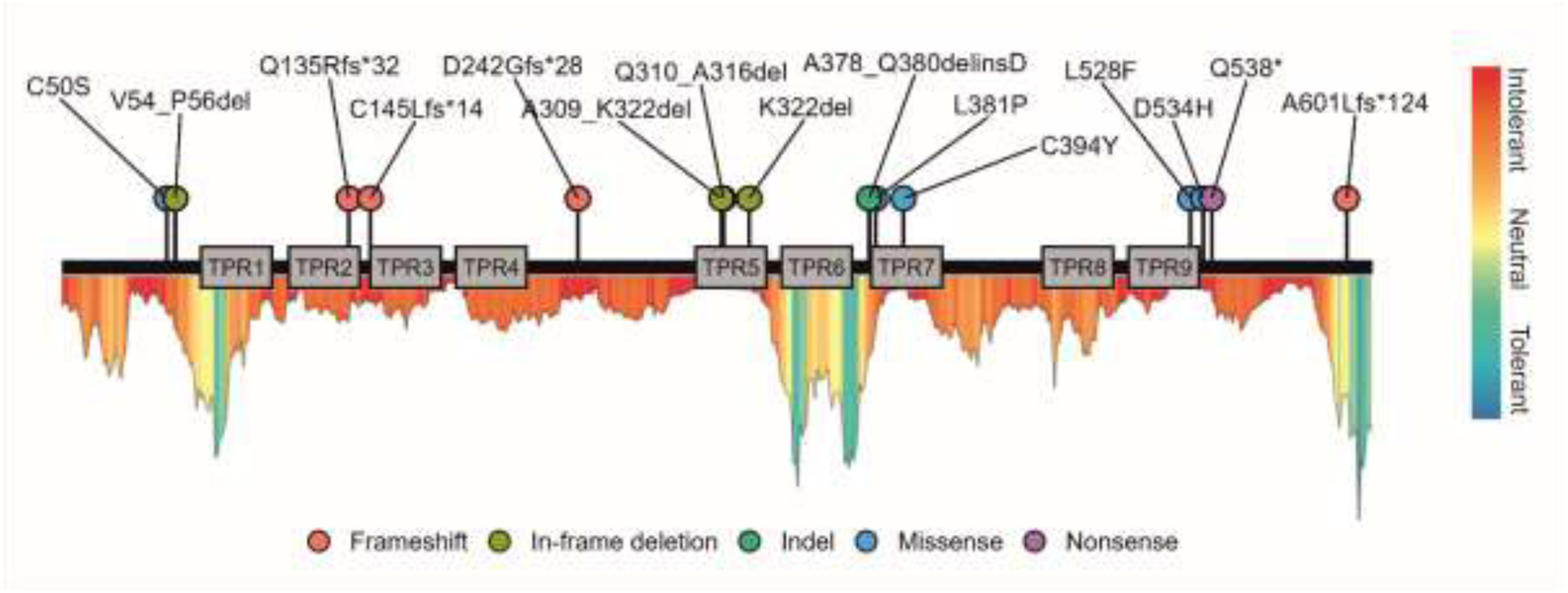
Schematic representation of the MAU2 protein and its tetratricopeptide repeat (TPR) repeats, with the relative positions of the variants identified in this study. Variant types are represented by colored lollipops: frameshift deletions/duplications (coral), in-frame deletions (olive green), delins (green), missense variants (blue), and nonsense variants (violet). A variation intolerance map derived from MetaDome (https://stuart.radboudumc.nl/metadome/) is also presented beneath the protein structure.

The amino acid residues affected by the missense substitutions and in-frame deletions map in regions with intolerance to variation, as evidenced by the residue-specific intolerance profiles generated by MetaDome^16^ (Figure 1). The potential impact of the missense variants was additionally assessed using six *in silico* prediction tools (Supplementary Table 1). Each variant was classified as deleterious by at least three of these tools. The identified variants occurred *de novo* in eight individuals (Patients 1, 3, 4, 5, 9, 10, 13, and 17) (Supplementary Tables 2 and 3). In five cases, assessment of inheritance was not possible due to the unavailability of one or both parents for genetic testing (Patients 2, 7, 8, 12, and 18). The remaining five variants were inherited from an affected parent exhibiting similar and/or milder clinical features. For Patients 11 and 16, sufficient clinical information was available for the affected carrier parents, who were therefore included in the cohort as Patients 12 and 17, respectively. The lack of sufficient clinical data for the carrier father of Patients 14 and 15, who are siblings, and of Patient 6, precluded the inclusion of these two parents in the cohort. Through application of the relevant ACMG criteria, variants were classified as pathogenic in four individuals, likely pathogenic in eleven, and of uncertain significance in three (Supplementary Table 2).

### Methylation profiling of *MAU2* patients reveals overlap with the CdLS episignature

Variants in four of the six primary CdLS-associated genes (*NIPBL*, *SMC1A*, *SMC3*, and *RAD21*) have been shown to result in overlapping DNA methylation profiles, collectively referred to as CdLS episignature^17^. This prompted us to investigate whether variants in *MAU2* might result in the same methylation pattern. To explore this, we analyzed blood-derived DNA from 13 individuals of our cohort, as well as from the carrier father of Patients 14 and 15.

EpiSign analysis revealed that ten cases exhibited a genome-wide DNA methylation profile similar to CdLS cases (Figures 2A, 2B, 2C). According to the EpiSign interpretation framework, a result is considered positive when at least two of the following criteria are met: (1) elevated methylation variant pathogenicity (MVP) score, (2) hierarchical clustering with positive controls, and (3) multidimensional scaling (MDS) proximity to reference cases. Although several samples had MVP scores below the typical threshold of 0.5 (Figure 2D), they were still classified as positive for the CdLS episignature because the other two analysis parameters (hierarchical clustering and MDS) matched the disorder-specific positive controls. The lower MVP scores observed in these cases may reflect a milder or variant-specific methylation profile, in which epigenetic alterations are present but less pronounced across the full CdLS episignature. Alternatively, this may be attributed to the fact that the canonical CdLS episignature was not originally defined using cases with *MAU2* variants.

**Figure 2.**
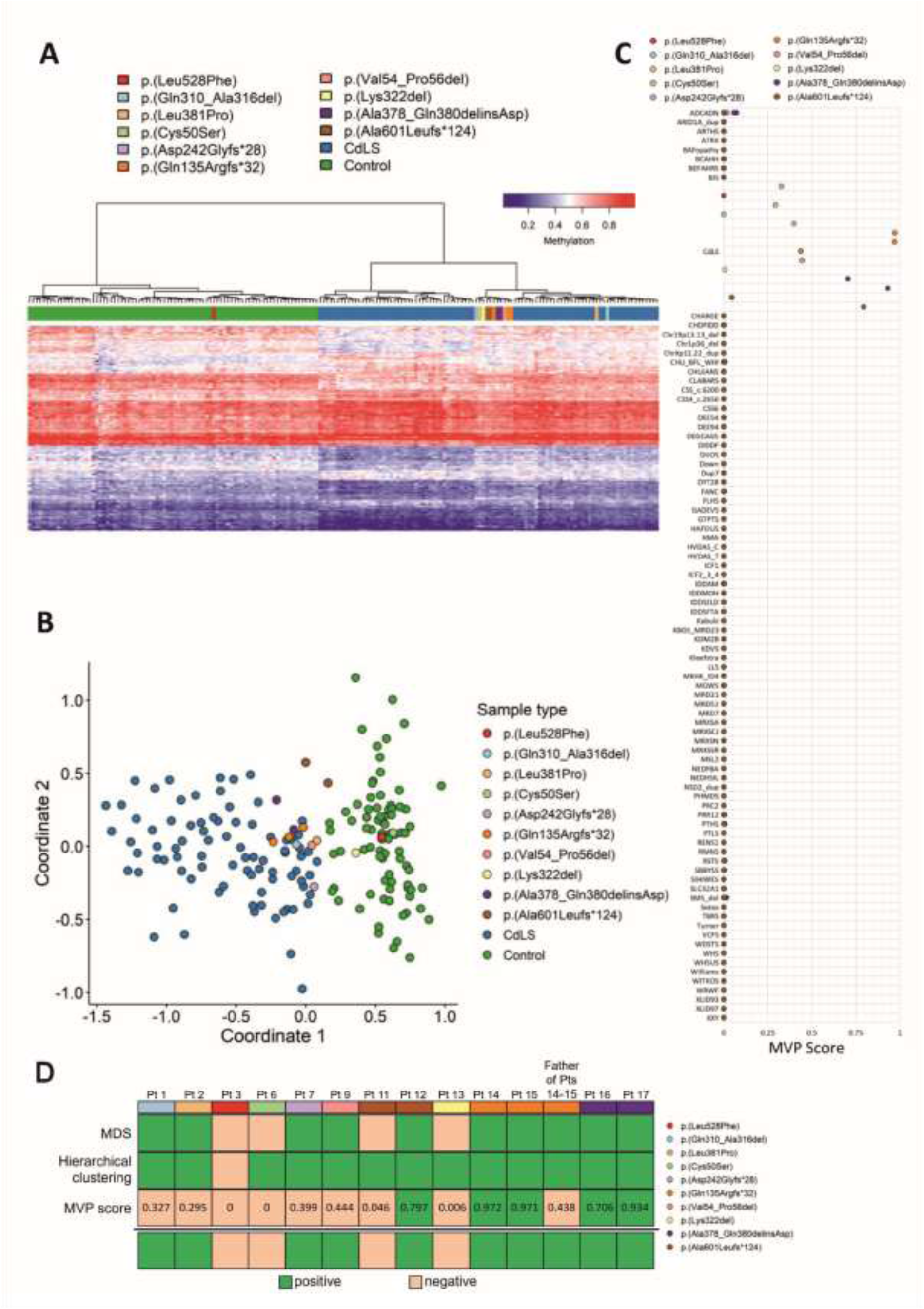
EpiSign (DNA methylation) analysis of peripheral blood from 14 individuals with 10 unique variants in *MAU2*. (A) Hierarchical clustering reveals that the *MAU2* cases exhibit a DNA methylation profile that is more similar to subjects with a confirmed CdLS episignature (blue), than with controls (green). One case, p.(Leu528Phe) (red) does not cluster with the rest of the cohort and shows a DNA methylation profile more similar to controls (green). (B) Multidimensional scaling analysis demonstrates that 10 cases (orange, purple, brown, pink, light orange, light blue, light purple) cluster with the confirmed CdLS cohort (blue) and are distinct from controls (green). In contrast, cases negative for the CdLS episignature (red, light green, yellow, brown) cluster with controls (green). (C) MVP score, a multi-class supervised classification system capable of discerning between multiple episignatures by generating a probability score for each episignature. 10 out of 14 cases (light blue, light orange, light purple, orange, pink, purple, brown) display elevated scores for CdLS, indicating a profile similar to the CdLS reference cohort. Conversely, four cases (red, light green, yellow, brown) show scores close to 0, suggesting a DNA methylation profile more similar to controls than to the CdLS cohort. (D) Summary of the DNA methylation analysis results with respect to the CdLS episignature.

In contrast, the four cases negative for the CdLS episignature (p.(Leu528Phe), p.(Cys50Ser), p.(Lys322del), and p.(Ala601Leufs*124)) exhibited methylation profiles closer to controls, consistent with their lower MVP scores. Three of these variants (p.(Cys50Ser), p.(Lys322del), and p.(Ala601Leufs124)) clustered with other *MAU2* cases on the CdLS side of the heatmap (Figure 2A), but grouped with controls in both the heatmap and MDS plot when analyzed separately (Supplementary Figure 1).

Since three of the four samples that did not align with the CdLS episignature clustered with CdLS cases when other *MAU2* samples were included in the hierarchical clustering analysis, we wondered whether there might be a *MAU2*-specific episignature. To address this, an episignature discovery analysis using *MAU2*-specific probes was performed. The analysis revealed two distinct *MAU2*-specific episignatures within the cohort, each consisting of a distinct but partially overlapping subset of *MAU2* samples (Supplementary Figures 2 and 3). The two *MAU2*-specific episignatures included both CdLS positive and CdLS negative cases. *MAU2* Episignature 1 comprised nine cases carrying six distinct variants: p.(Cys50Ser), p.(Asp242Glyfs28), three individuals with p.(Gln135Argfs32), p.(Val54_Pro56del), p.(Lys322del), and two individuals with p.(Ala378_Gln380delinsAsp) (Supplementary Fig. 2). *MAU2* Episignature 2 included seven cases with three distinct variants: three with p.(Gln135Argfs32), two with p.(Ala378_Gln380delinsAsp), and two with p.(Ala601Leufs124) (Supplementary Fig. 3). Within each *MAU2* episignature, a subset of similarly differentially methylated probes exhibited consistent methylation across the included cases, allowing each episignature to be defined. Some variants contributed to both episignatures, others to one or neither, highlighting heterogeneity in *MAU2* methylation profiles. Strikingly, in a mother–son pair with the p.(Ala601Leufs*124) variant (Patients 11 and 12), CdLS episignature results were discordant: the mother tested positive, while the son did not, despite retaining the *MAU2* episignature 1. The son’s sample also showed an epigenetic age mismatch (chronological 16 years, predicted 27 years), suggesting that technical or biological factors may have affected detection of the CdLS-specific episignature in this patient.

The subset of similarly differentially methylated CpGs defining each episignature reflects shared methylation features among the included cases, while each episignature captures only a portion of the overall global methylation profile. These episignature groupings may reflect biological factors such as variant-specific effects, differences in variant location, similar functional consequences, or variability in clinical phenotype.

### A subset of *MAU2* variants impairs NIPBL–MAU2 heterodimerization

MAU2 promotes proper folding and stability of NIPBL by enwrapping its N-terminal region^9^. *MAU2* variants that affect residues critical for this molecular interface may therefore compromise the integrity of the MAU2-NIPBL interaction. Supporting this hypothesis, we have shown that the previously reported MAU2 p.(Gln310_Ala316del) in-frame deletion of Patient 1 impairs this interaction^12^. Expanding on this observation, we sought to systematically assess the impact of all missense substitutions and in-frame deletions of our cohort using a quantitative mammalian two-hybrid assay, as previously described^12^. This analysis revealed that six out of ten tested *MAU2* variants significantly impaired complex formation (Figure 3). Three in-frame deletions partially overlapped within the same region of MAU2: p.(Ala309_Lys322del), p.(Gln310_Ala316del), and p.(Lys322del). The two deletions spanning multiple amino acids markedly disrupted the interaction, whereas the p.(Lys322del) affecting the terminal end of this region had a milder, non-significant effect, indicating that both the position and extent of the deletion influence NIPBL binding efficiency. Importantly, the results of the mammalian two-hybrid assay were consistent with those of the episignature study in patients for whom blood DNA was available (Figure 3). Blood samples from patients with variants that significantly disrupted the MAU2-NIPBL interaction also showed a positive CdLS episignature (p.(Val54_Pro56del), p.(Gln310_Ala316del), p.(Ala378_Gln380delinsAsp), and p.(Leu381Pro)). Conversely, variants without significant interaction impairment (p.(Cys50Ser), p.(Lys322del), p.(Leu528Phe)) were negative for the CdLS episignature. Notably, two of these negative cases (p.(Cys50Ser) and p.(Lys322del)) displayed a positive *MAU2* episignature, suggesting that disruption of the NIPBL–MAU2 interaction is more closely tied to the CdLS molecular phenotype, whereas isolated *MAU2* episignatures may reflect subtler effects or MAU2-specific mechanisms independent of NIPBL.

**Figure 3.**
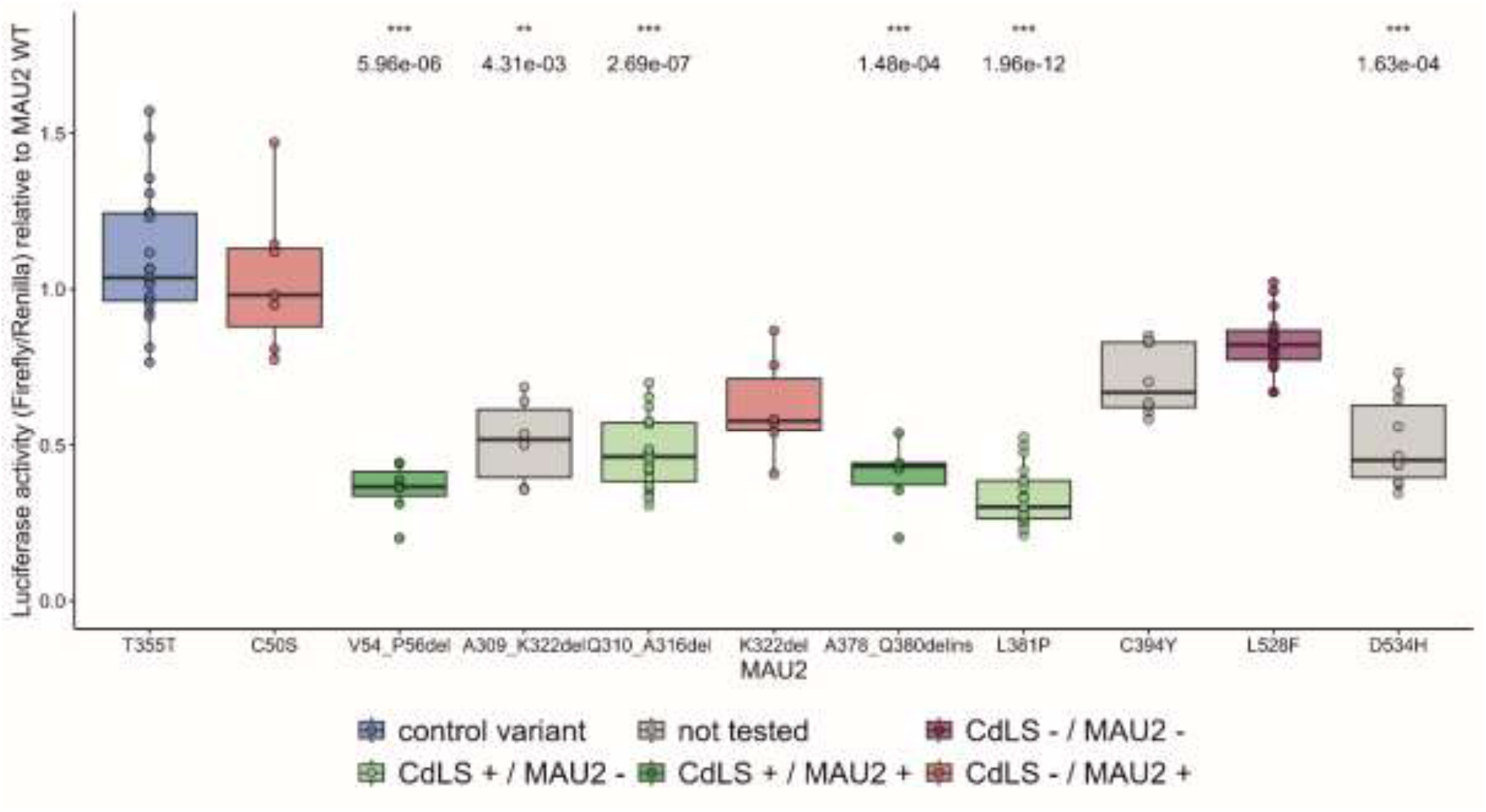
Results of the mammalian two-hybrid assay quantifying the interaction strength between MAU2 and NIPBL. The luciferase-to-Renilla ratio for each variant was normalized to that of the wild-type MAU2 construct, which was assigned a value of 1 in the graph. A control variant, T355T (reported as frequent in the healthy population: 625/1,612,168 alleles in gnomAD v4.1.0), was included as a negative control. The control variant T355T is highlighted in blue. Statistically significant *p*-values are indicated above the corresponding variant; variants lacking *p*-values did not achieve statistical significance (error bars +/− S.D., Dunn’s test followed by Bonferroni Correction for multiple comparisons, * *p* ≤ 0.05, ** *p* ≤ 0.01, *** *p* ≤ 0.001). The mammalian two-hybrid assay results were correlated with the episignature analysis. Samples not available for episignature testing are represented in grey, those exhibiting a positive CdLS episignature and a positive *MAU2* episignature in green, those exhibiting a positive CdLS episignature and a negative *MAU2* episignature in light green, those exhibiting a negative CdLS episignature and a positive *MAU2* episignature in red, and those with a methylation profile negative for both the CdLS and *MAU2* episignature in purple.

To distinguish whether the reduced signal in the mammalian two-hybrid assay resulted from impaired protein–protein interaction or from variant-induced MAU2-destabilization unrelated to the interaction, we evaluated the expression of all pCMV-AD_MAU2 constructs used in the assay by Western blotting using both MAU2- and NF-κB-specific antibodies, with GAPDH used as a loading control (Supplementary Figures 4A-B). However, as transfection efficiency could not be controlled for, the signal intensity of the MAU2 constructs was not normalized to GAPDH. One mutant, p.(Ala309_Lys322del), consistently showed reduced expression across replicates with both antibodies, suggesting possible protein instability in addition to impaired NIPBL interaction. A smaller deletion within the same region (p.(Gln310_Ala316del)), which does not appear to compromise protein stability, also disrupts the interaction. This supports the hypothesis that the reduced relative signal observed in the mammalian two-hybrid assay for the larger deletion (p.(Ala309_Lys322del)) is at least partially attributable to impaired NIPBL interaction. Interestingly, when expressed from an alternative 3xFLAG vector, p.(Ala309_Lys322del) levels were comparable to those of the other variants (Supplementary Figure 4C), indicating that vector-specific effects may also contribute the reduced signal observed with the pCMV-AD construct. Thus, both plasmid context and variant-specific properties may influence expression, a factor that should be considered when interpreting the mammalian two-hybrid results for this variant.

### Fibroblasts with a frameshift variant exhibit *MAU2* haploinsufficiency

The aforementioned experiments have shown that the majority of *MAU2* in-frame variants impair the interaction with NIPBL. However, the functional consequences of truncating variants still remain to be elucidated. To explore this, we analyzed primary dermal fibroblasts from carriers of the frameshift deletion p.(Gln135Argfs*32). As cells from Patient 15 failed to expand, experiments were performed on cells of Patient 14 and of her carrier father. These samples were used to assess the impact of the frameshift variant on *MAU2* transcript levels by qPCR. The analysis was extended to include Patient 1, who carries an in-frame deletion known to disrupt the MAU2–NIPBL interaction. *MAU2* transcript levels were normalized to two independent housekeeping genes (*NADH* and *GAPDH*) and compared to the mean expression levels of five unrelated healthy controls. A total of six independent experiments were conducted across three biological replicates (two technical replicates per RNA preparation). The results revealed a statistically significant reduction of the *MAU2* transcript levels in both Patient 14 and her father, consistently across both normalization strategies (Figures 4A and 4B, Supplementary Figure 5). In contrast, *MAU2* transcript levels in fibroblasts from Patient 1 were comparable to control levels. This indicates that the frameshift deletion p.(Gln135Argfs*32), but not the in-frame variant p.(Gln310_Ala316del), leads to reduced *MAU2* transcript abundance, supporting nonsense-mediated mRNA decay (NMD)–induced haploinsufficiency as the likely pathogenic mechanism. We next analyzed *NIPBL* transcript levels, which showed no consistent changes across samples. A statistically significant reduction was observed in Patient 14 when normalized to *GAPDH* only (Figures 4C and 4D), likely reflecting housekeeping gene variability rather than true downregulation, as the patient’s father showed no changes in *NIPBL* expression levels. These observations indicate that the apparent reduction in Patient 14 is a normalization artifact, and overall, that there is no evidence of a transcriptional feedback regulating *NIPBL* expression in response to *MAU2* haploinsufficiency. Although *NIPBL* transcript levels were unaffected, a reduction of NIPBL at the protein level may still occur due to the mutual dependence between NIPBL and MAU2 for stability. The analysis of the total lysates by Western Blot confirmed that both MAU2 and NIPBL proteins are less abundant in Patient 14 and her father (Figure 4E, Supplementary Figure 6), demonstrating that MAU2 haploinsufficiency extends to NIPBL reduction at the protein level.

**Figure 4.**
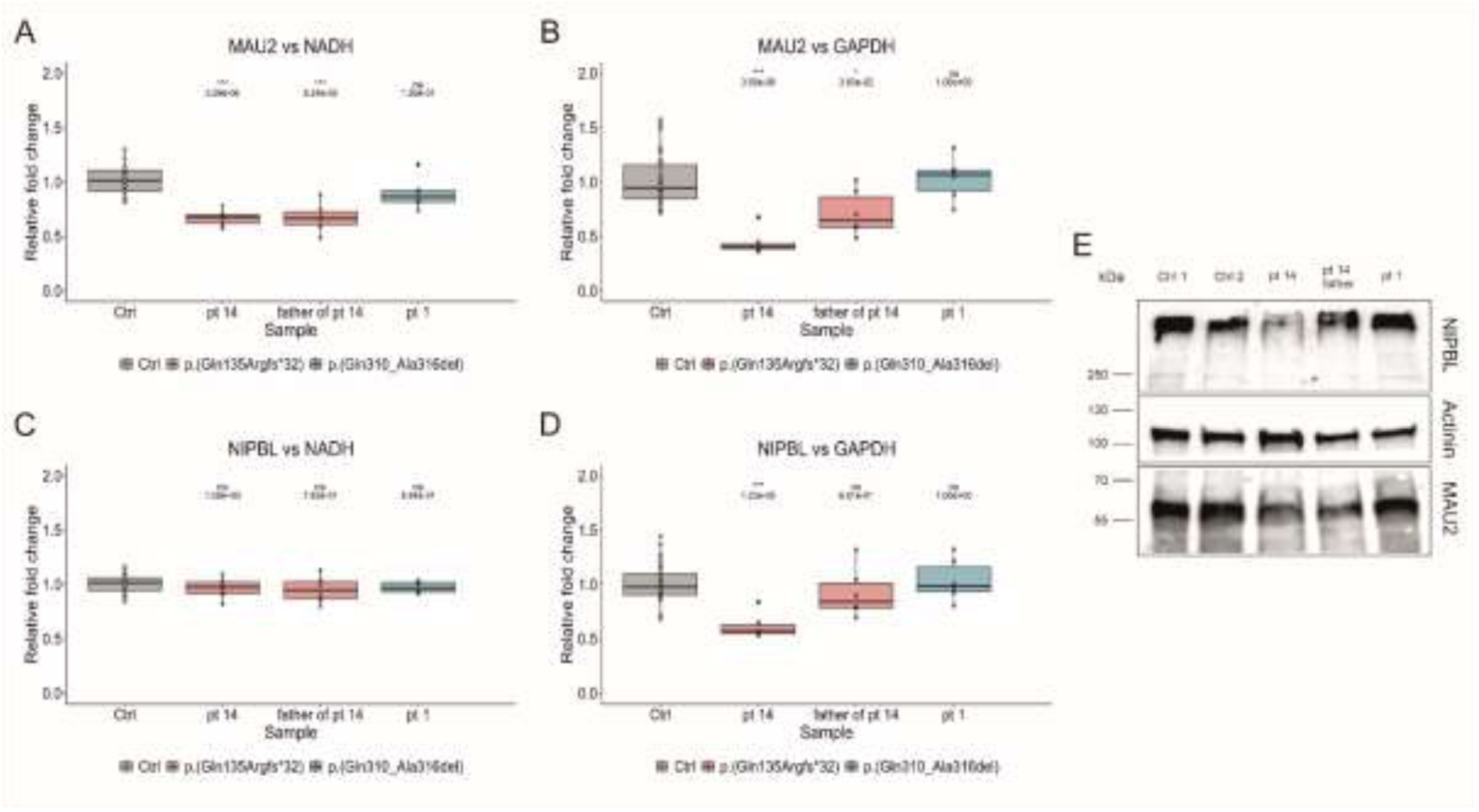
Fibroblasts with a frameshift variant exhibit MAU2 haploinsufficiency. (A-D) Quantification of *MAU2* and *NIPBL* transcript levels by qPCR. In all panels, data from five unrelated healthy controls are represented in grey, samples harboring the frameshift variant p.(Gln135Argfs*32) are shown in red, and Patient 1 with the in-frame variant known to disrupt NIPBL interaction (p.(Gln310_Ala316del)) is shown in blue. Six independent experiments were performed, with two technical replicates per RNA preparation (error bars +/− S.D., Mann-Whitney U test followed by Bonferroni Correction for multiple comparisons, * *p* ≤ 0.05, ** *p* ≤ 0.01, *** *p* ≤ 0.001). (A) *MAU2* transcript levels normalized to *NADH*. (B) *MAU2* transcript levels normalized to *GAPDH*. (C) *NIPBL* transcript levels normalized to *NADH*. (D) *NIPBL* transcript levels normalized to *GAPDH*. (E) Quantification of MAU2 and NIPBL protein levels by Western Blot. Protein levels were assessed in two healthy controls (Ctrl 1 and Ctrl 2), Patient 14 and her father (*MAU2* frameshift variant), and in Patient 1 (*MAU2* in-frame deletion). The membranes were cut to allow all three proteins (NIPBL, MAU2, and the loading control Actinin) to be visualized from a single blot.

### Variants in *MAU2* result in variable phenotypes mainly characterized by short stature and microcephaly

Our cohort of patients with *MAU2* variants comprises nine males and nine females, ranging in age from 20 months to 39 years (median age 12 years). Comprehensive phenotypic data for each individual are provided in Supplementary Table 3. The core clinical features observed in our cohort are summarized in Table 1. The most frequently reported phenotypes include short stature (83%), microcephaly (75%), and intellectual disability (61%) (Figure 5A). Additional recurrent findings were synophrys (56%), long philtrum (56%), thick eyebrows (50%), smooth philtrum (50%), and a thin upper lip vermilion (44%). Central nervous system anomalies were present in 42% of individuals, anteverted nostrils in 39%, behavioral abnormalities in 38%, and hypertrichosis in 38%. Speech delay was reported in 35% of the cases, genitourinary anomalies in 27%, and downturned corners of the mouth in 22% (Figure 5A). These observations indicate that short stature and microcephaly are the most consistently observed clinical features associated with *MAU2* variants. Both were also often present at birth, with microcephaly and short stature observed in approximately 50% of individuals for whom neonatal data were available (5/11 and 8/16, respectively). One third of patients with short stature received growth hormone (GH) treatment (Patients 2, 5, 14, 15, and 17) (Supplementary Table 3). The treatment began as early as 1 month and as late as 13 years. Although several patients were treated for up to ten years, improvement in height trajectory was sporadic and not observed consistently across all individuals. Intellectual disability (ID) was the third most common feature, although it varied in severity among patients. The majority exhibited learning difficulties (4/11) or borderline-to-mild ID (4/11), while only three patients showed moderate-to-severe ID (Figure 5B). Thus, cognitive impairment in these patients appears to be generally mild.

**Figure 5.**
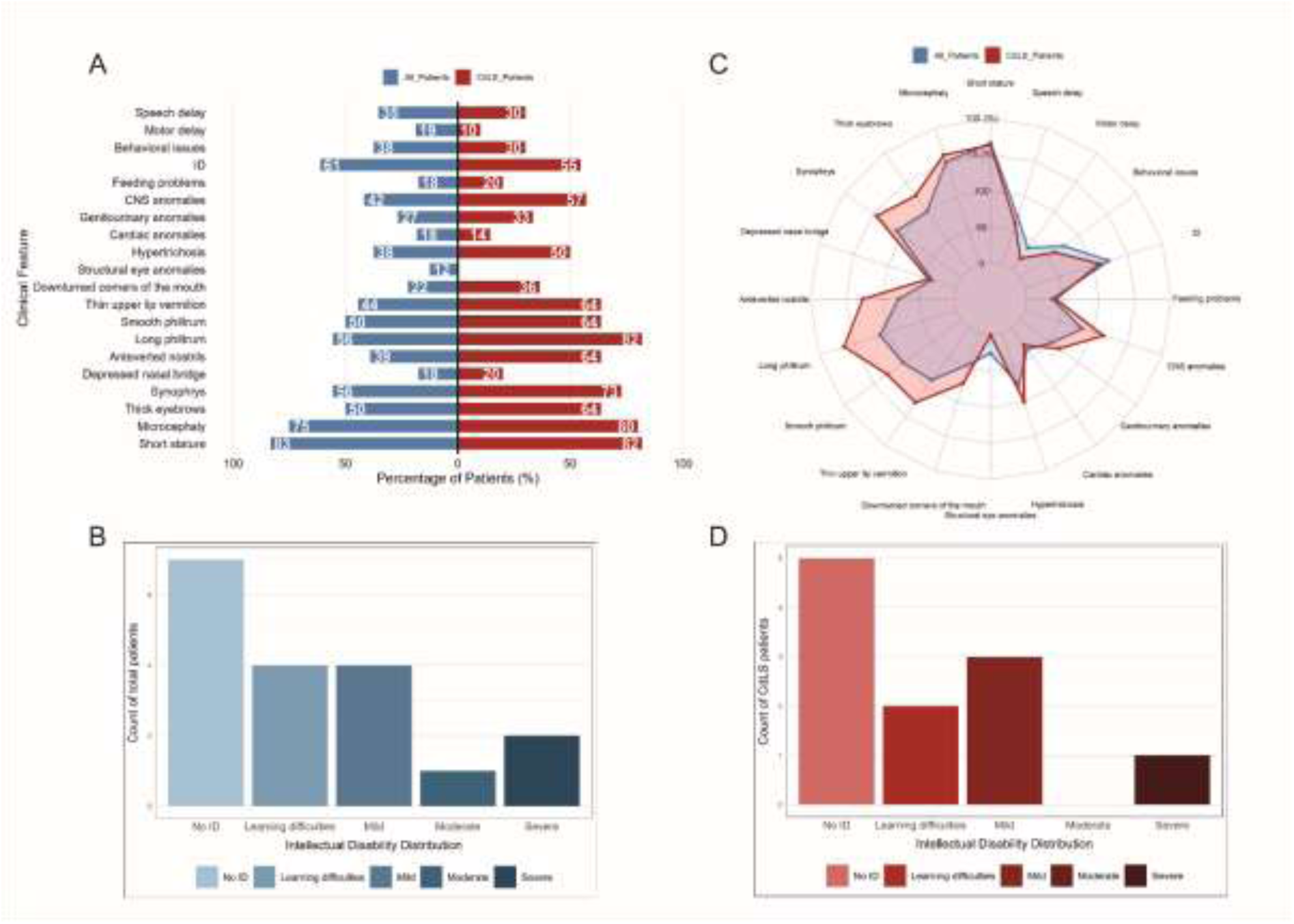
Frequency of key clinical features associated with *MAU2* variants. (A) Mirror plot illustrating the frequency (as a percentage) of the main clinical features in the total cohort (blue) compared to molecularly confirmed CdLS patients (red). (B) Distribution of intellectual disability (ID) severity within the total cohort. (C) Radar plot emphasizing the increased prevalence of facial dysmorphism in molecularly confirmed CdLS patients. (D) Distribution of ID severity within the molecularly confirmed CdLS cohort.

**Table 1.**
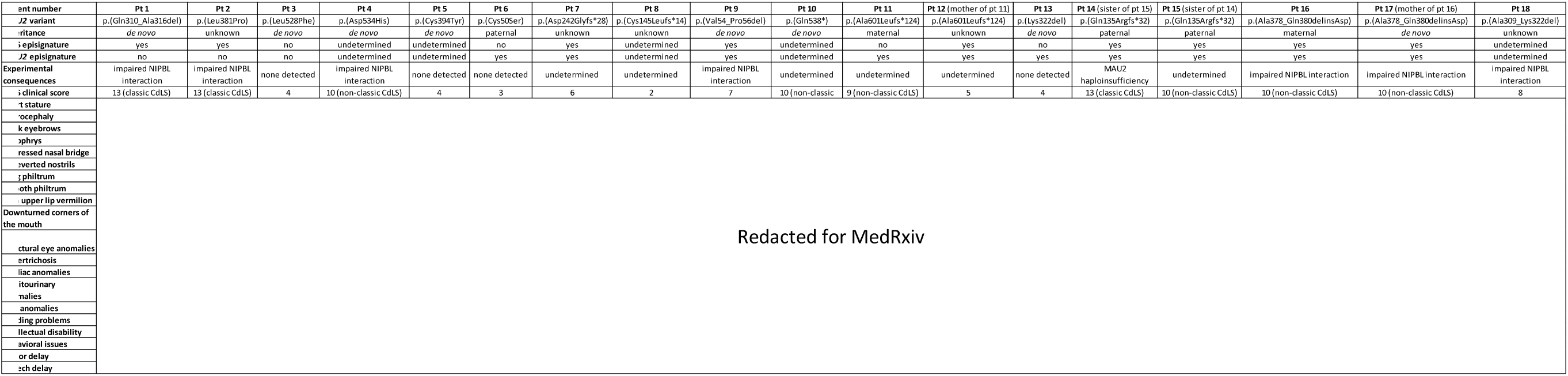
Summary of the clinical features and experimental results.

Behavioral anomalies were reported in fewer than half of the individuals, with attention-deficit/hyperactivity disorder (ADHD), hyperactivity, and anxiety being the most common features. Developmental milestones were achieved in the majority of cases within the expected age range, with a median age of independent walking at 15 months and first spoken words at 14 months. Brain MRI abnormalities were also frequently observed, although the findings were largely nonspecific.

Among the 18 patients, molecular evidence supporting a diagnosis of CdLS was obtained in 11 cases (Table 1), either via a positive CdLS episignature or significant disruption of NIPBL–MAU2 interaction. Three additional patients showed only a *MAU2*-specific episignature, suggesting a milder or variant-specific effect, while two tested negative across all molecular assays conducted in this study. Appropriate testing was not possible for the remaining two patients due to sample unavailability (Table 1). To explore whether patients with molecularly confirmed CdLS exhibit a recognizable clinical profile, we compared the frequency of clinical features observed across the entire cohort with that in the subset of confirmed CdLS cases (defined by a positive CdLS episignature or impaired NIPBL–MAU2 interaction). While microcephaly and short stature were comparably frequent across both groups, there was a higher frequency of facial dysmorphism among confirmed CdLS patients (Figures 5A and 5C). The features with a higher frequency in the CdLS-subgroup included thick eyebrows, synophrys, anteverted nostrils, a long and smooth philtrum, thin upper lip vermilion, and downturned corners of the mouth. These characteristic facial features can be recognized in the facial photographs of the individuals who consented to publication (Figure 6). The distribution of ID severity was instead comparable to that observed in the full cohort (Figure 5B and 5D).

**Figure 6.** Facial appearance of *MAU2* patients.

Given the association between *MAU2* and CdLS emerging from our data, we applied the published CdLS clinical scoring system to all individuals in the cohort (Table 1, Supplementary Table 4). Three individuals met criteria for classic CdLS, each scoring 13 points (Patients 1, 2, and 14). Six individuals fell within the non-classic CdLS range (9–10 points) (Patients 4, 10, 11, 15, 16, and 17), while five scored between 4 and 8 points and presented with at least one cardinal feature, thus meeting the criteria for molecular testing for CdLS (Patients 3, 7, 9, 13, and 18). The remaining four individuals scored below 4, or above 4 without any cardinal features, thus falling outside the diagnostic threshold for CdLS (Patients 5, 6, 8, and 12). Of note, intrafamilial differences were observed in some cases. For instance, whereas patients 16 and 17 (index patient and his mother) both scored 10, Patient 11 scored 9, while his mother (Patient 12) scored 5; Patient 14 scored 13, whereas her sister (Patient 15) scored 10. Remarkably, all three individuals classified as classic CdLS belonged to the molecularly confirmed CdLS group. Among the six individuals with non-classic CdLS scores, four had molecular confirmation (Patients 4, 15, 16, and 17). Of the remaining two, one exhibited a *MAU2*-specific episignature (Patient 11), while the other could not be tested due to unavailability of samples (Patient 10). On the other hand, none of the two individuals with negative results in both episignature profiling and protein interaction assays (Patients 3 and 5) exceeded the diagnostic threshold for CdLS (≥5 points). This concordance between clinical scoring and molecular data supports the reliability and specificity of the molecular assays used in this study.

### Heterozygous *Mau2* knockout mice display decreased body length, microcephaly, and brain structural anomalies

To investigate whether the consequences of *Mau2* loss-of-function *in vivo* mirror the phenotypic features observed in *MAU2* patients, we generated a constitutive *Mau2* knockout mouse line in collaboration with the Sanger Institute Mouse Genetics Project^18^. The *Mau2 ^tm1b(KOMP)Wtsi^*mice were generated using the knockout-first allele method^19^ developed by the International Mouse Phenotyping Consortium (IMPC), in which exon 5 of *Mau2* is floxed and a LacZ cassette inserted (Figure 7A).

**Figure 7.**
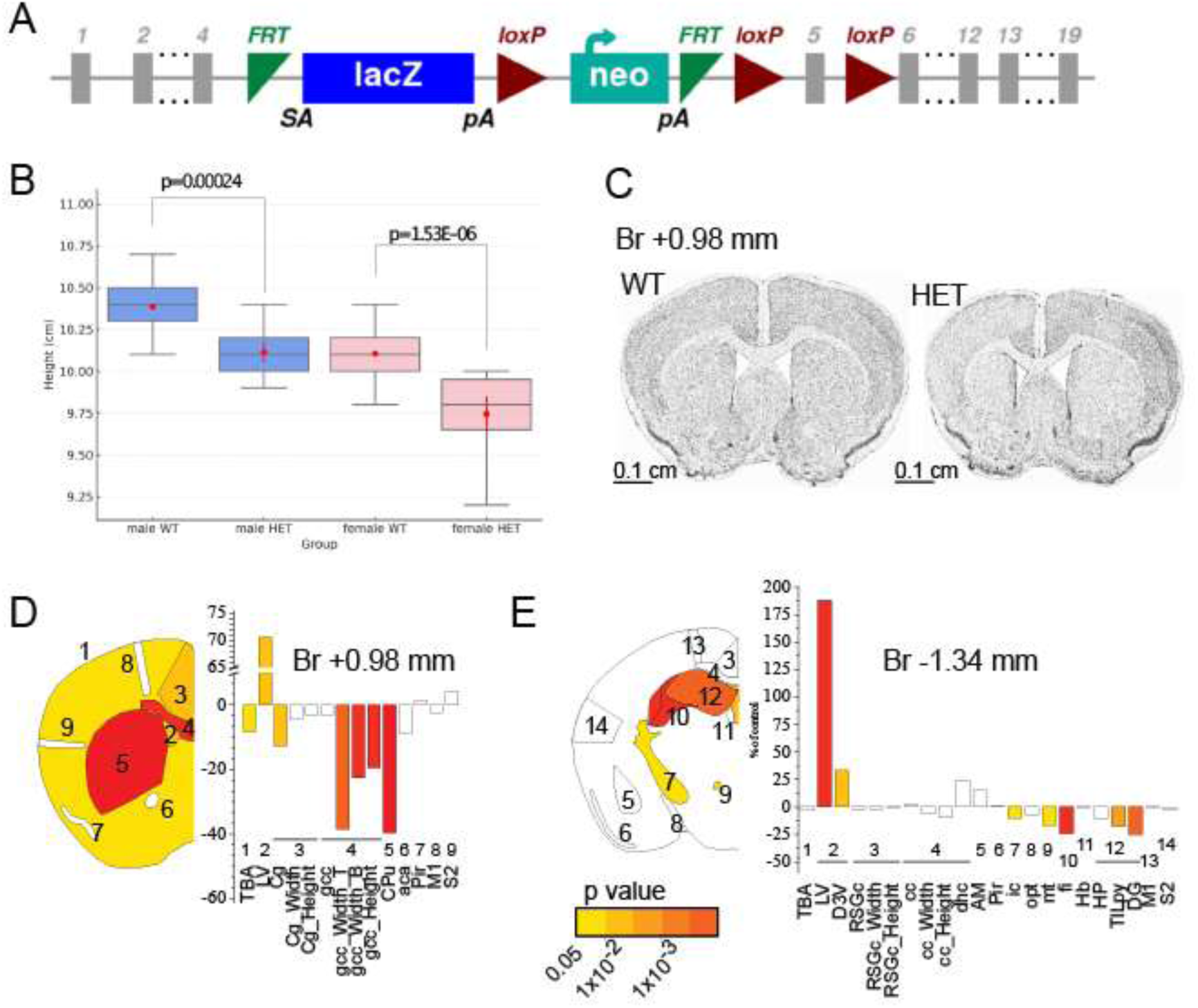
*Mau2* haploinsufficiency leads to microcephaly and region-specific neuroanatomical defects in the *Mau2*^+/−^ mouse. (A) Tm1a genetic construct showing the insertion of the LacZ and Neo cassettes upstream the critical exon 5 of *Mau2*. (B) Body length boxplots showing the median (center line), interquartile range (box), and range excluding outliers (whiskers). Individual data points are overlaid (black dots). Males are shown in blue, females in pink. Red markers represent the group mean ± standard error of the mean (SEM). Groups are displayed as: Male WT (n=345), Male HET (n=7), Female WT (n=340), Female HET (n=7). (C) Representative coronal brain sections from *Mau2^tm1b^* mouse (left) and matched wildtype (right), showing the microcephaly phenotype at Bregma (Br) +0.98 mm. Scale bar: 0.1 cm. (D) Schematic representation of affected brain regions at Bregma +0.98 mm according to p-values in three adult male *Mau2*^+/−^ mice aged 16 weeks compared to matched wild-type mice (n=44). Histograms represent percentage change relative to WT mice (set as 0) for each of the measured parameters. Numbers indicate a group of studied brain regions. Brain structures assessed: total brain area (TBA); lateral ventricles (LV); cingulate cortex (Cg); genu of the corpus callosum (gcc); caudate putamen (CPu); anterior commissure (aca); piriform cortex (Pir); primary motor cortex (M1); secondary somatosensory cortex (S2). Data were analyzed by two-tailed Student’s *t*-tests assuming equal variances. (E) Schematic representation of affected brain regions at Bregma −1.34 mm, analysed as in (D). Histograms represent percentage change relative to WT. Brain structures assessed: the total brain area (TBA); lateral- (LV) and third ventricles (D3V); retrosplenial granular cortex (RSGc); corpus callosum (cc); dorsal hippocampal commissure (dhc); amygdala (AM); piriform cortex (Pir); internal capsule (ic); optic tract (opt); mammillothalamic tract (mt); fimbria (fi); habenular nucleus (Hb); hippocampus (HP); total internal length of pyramidal cells (TILpy); dentate gyrus (DG); primary motor cortex (M1); secondary somatosensory cortex (S2). The color code indicates the significance threshold, or white when not significant and grey when not computable. Data were analyzed by two-tailed Student’s *t*-tests of equal variances.

Mouse survival was assessed from 102 successfully genotyped mice derived from a heterozygous-by-heterozygous (“het-by-het”) breeding scheme. The observed genotypic distribution was 0% homozygous, 57% heterozygous, and 43% wildtype (Supplementary Table 5), indicating that the complete loss of *Mau2* is incompatible with life, consistent with a previous report^20^. Since only heterozygous mice could be generated, the *Mau2 ^tm1b(KOMP)Wtsi^* mouse model will be hereafter referred to as *Mau2*^+/−^. However, we recovered more heterozygous females than males (37 vs. 21), and this sex imbalance suggests a previously unreported sub-viability associated with the heterozygous state that is specific to males (Supplementary Table 5). A one-tailed binomial test confirmed that the excess of females was significant (*p*=0.024).

Surviving mice were assessed for their body weight and body length (from the nose to the beginning of the tail) at 16 weeks of age (Supplementary Table 6). Although no significant difference was found in body weight, the length of the mice was consistently reduced by 4% in female heterozygous (*p*=1.53E-06) and by 3% in male heterozygous (*p*=0.00024) mice (Figure 7B).

We next assessed brain anatomy in 16-week-old *Mau2*^+/−^ mice using a standardized high-resolution pipeline that measures 36 unique parameters across 23 brain regions (Supplementary Table 7), following our previously established protocols^21, 22, 23^. At Bregma +0.98 mm, the brain size in *Mau2*^+/−^mice was reduced by 10% (*p*=0.0168), indicative of moderate microcephaly (Figure 7C). We also noted a clear left-right asymmetry of the brain, particularly evident in the caudate putamen and lateral ventricles at Bregma +0.98 mm (Figure 7C). Compared to wildtype mice, *Mau2*^+/−^ mice exhibited a total of ten significantly altered brain regions. At Bregma +0.98 mm, these included the lateral ventricles, the cingulate cortex, the caudate putamen, the genu of the corpus callosum, and the total brain area (Figure 7D). At Bregma −1.34 mm, significant changes were observed in the internal capsule, the hippocampal formation, the dorsal third ventricle, the mammillothalamic tract, and the fimbria of the hippocampus (Figure 7E). While the ventricles showed enlargement, all other affected brain regions were reduced in size. For example, the cingulate cortex was reduced by 13% (*p*=0.0065), the internal capsule by 10.1% (*p*=0.026), the total length of pyramidal cell layer by 17.5% (*p*=0.0018), and the height of the genu of the corpus callosum by 19.5% (*p*=4.72E-05) (Figure 7D-E, Supplementary Table 8). These analyses indicate that the microcephaly phenotype does not reflect equally small brain regions, but instead represents a mosaic pattern of region-specific changes. Taken together, *Mau2*^+/−^ heterozygous knockout mice exhibit short stature, microcephaly, and region-specific neuroanatomical anomalies, recapitulating key clinical features of *MAU2*-associated phenotypes observed in patients.

## Discussion

Before this study, the disease-causing potential of *MAU2* variants was unclear. Only a few cases had been reported, including two *de novo* in-frame variants and three *de novo* microdeletions at 19p13.11–p12 associated with broad, nonspecific neurodevelopmental phenotypes^12, 13, 14^. The large size of these deletions (1.4–5.2 Mb) and the involvement of numerous additional genes (37–68 genes) precluded definitive attribution of the observed phenotypes to *MAU2* alone. Moreover, the scarcity of reported variants had limited our understanding of the phenotypic spectrum and underlying pathogenic mechanisms connected to *MAU2* dysfunction. Our characterization of 18 individuals with 15 distinct *MAU2* variants directly addresses these knowledge gaps, providing robust evidence for its pathogenic role and advancing our understanding of the associated clinical features.

To explore the molecular consequences of *MAU2* loss-of-function variants, we analyzed patient-derived fibroblasts carrying a frameshift variant. In these cells, *MAU2* mRNA levels were significantly reduced, likely resulting from degradation of the mutant transcript via nonsense-mediated decay (NMD). This supports a haploinsufficiency mechanism at the RNA level for *MAU2* loss-of-function variants, similar to what has been previously observed for *NIPBL* variants^24^. In contrast, *NIPBL* transcript levels remained unaffected in the *MAU2* mutant cells, indicating the absence of a direct transcriptional feedback regulation between the two cohesin loader subunits. However, protein analysis revealed reduced levels of both MAU2 and NIPBL, consistent with prior siRNA-based studies showing that loss of one cohesin loader subunit destabilizes the other^10^. Similar effects were observed in induced pluripotent stem cells with *NIPBL* variants^25^. Although our analysis was limited to a single family, it is plausible that other loss-of-function *MAU2* variants might similarly trigger NMD and result in *MAU2* haploinsufficiency.

Given the interdependence between MAU2 and NIPBL, in-frame *MAU2* variants that weaken this interaction would be expected to similarly reduce protein stability of both MAU2 and NIPBL. However, this anticipated effect was not observed in our experiments: despite substantial impairment of the interaction, protein levels remained stable in fibroblasts carrying an in-frame deletion. This may reflect a combination of dosage sensitivity and compensatory mechanisms. Functional assays showed that in-frame variants preserve partial MAU2–NIPBL binding *in vitro* (at least 30%), allowing some complex formation. In addition, as previously reported for *NIPBL* variants, upregulation of the wild-type allele may further compensate, preventing major reductions in protein levels^24^. Together, these factors likely result in a greater amount of functional MAU2 protein in cells with in-frame variants compared to variants leading to *MAU2* haploinsufficiency. Moreover, even a weakened interaction may be sufficient to maintain the stability of NIPBL and MAU2 and prevent their degradation, although it may not fully sustain downstream cohesin loader functions, such as interaction with additional transcription factors. Reflecting a shared pathogenic mechanism characterized by partial impairment of cohesin loader function, both in-frame and truncating *MAU2* variants lead to a DNA methylation profile that overlaps with the established CdLS episignature^17, 26, 27, 28^, providing strong molecular evidence linking *MAU2* variants to CdLS. Importantly, we were also able to establish a novel *MAU2*-specific episignature. Interestingly, some patients do not display the CdLS episignature but are positive for the *MAU2* episignature, indicating that certain *MAU2* variants may partially impair cohesin loader function in a way that is sufficient to generate a *MAU2*-specific methylation signature, yet insufficient to trigger the full CdLS DNA methylation profile.

In our cohort, the most common phenotypic features associated with *MAU2* variants were short stature and microcephaly, often present from birth. One-third of patients with short stature (5/15) received growth hormone (GH) treatment. GH deficiency was confirmed in two cases, absent in one, and unknown in the remaining patients (Supplementary Table 3). Treatment was initiated between 1 month and 13 years of age, with durations up to 10 years, but generally yielded limited benefit. A direct comparison between two siblings highlighted a possible potential benefit: Patient 15, who had GH deficiency and started treatment earlier than her sister (Patient 14, who does not present with GH deficiency), demonstrated a milder degree of short stature (−2.02 SD *vs*. −2.97 SD) and her height improved from −2.29 to −2.02 SD during the last year of treatment. GH treatment remains a largely unexplored area, even among individuals with CdLS not associated with *MAU2* variants. GH secretion is typically within the normal range in most children with CdLS. Evidence suggests that, even in the absence of GH deficiency, patients with short stature may derive modest benefit from GH therapy^29^. In one previously reported case, a CdLS patient without evidence of GH deficiency or insensitivity initiated GH treatment at 4.3 years of age and achieved a substantial height gain of 1.6 SD over eight years, with no significant adverse reaction^30^. Given the limited number of treated individuals, GH therapy may be considered on a case-by-case basis in *MAU2* patients with significant short stature, particularly if GH deficiency is documented, but its efficacy remains uncertain.

Most individuals with *MAU2* variants achieved developmental milestones within the expected age range, in contrast to the more pronounced delays typically observed in CdLS patients with *NIPBL* variants^1^. Furthermore, ID was generally milder in the *MAU2* group compared to non-*MAU2* CdLS cases^1^, and some individuals in our cohort did not display any type of cognitive impairment.

Clinical expressivity appears to be variable among individuals with *MAU2* variants, as illustrated by the few familial cases in our cohort. Nevertheless, penetrance seems to be complete, since all carrier parents displayed at least some clinical features observed in their affected children. A particularly informative example involves two sisters (Patients 14 and 15) carrying a paternally inherited *MAU2* frameshift deletion. One sister presented with a CdLS score of 13, consistent with a classic phenotype, whereas the other scored 10. This difference likely reflects milder or absent facial dysmorphism in the younger sibling. Despite both sisters meeting criteria for either classic or non-classic CdLS based on their scores, neither exhibited developmental delay or intellectual disability, highlighting that cognitive deficits in *MAU2*-related CdLS are milder than in *NIPBL*-related cases. These observations raise the question of whether developmental delay and intellectual disability might be underpowered in the current CdLS scoring system, as their combined contribution accounts for only a single point of the total score. More broadly, the CdLS score places limited emphasis on neurological manifestations, with seizures, behavioral anomalies, and neuropathological findings carrying limited weight on the total score.

Animal models have been instrumental in elucidating the developmental mechanisms underlying CdLS. In mice, *Nipbl* haploinsufficiency (*Nipbl*^+/−^) produces growth restriction, craniofacial anomalies, and neurodevelopmental defects that closely parallel human CdLS phenotypes^31, 32^. Complete systemic loss of either *Nipbl* or *Mau2* results in embryonic lethality before E9.5, underscoring the essential function of the cohesin loader complex in early development. To circumvent this limitation, Smith and colleagues generated conditional alleles^20^. Neural crest–specific inactivation of *Mau2* resulted in severe craniofacial hypoplasia, in some cases more pronounced than with *Nipbl* deletion. Notably, double mutants (*Nipbl*^−/−^; *Mau2*^−/−^) showed milder phenotypes than *Mau2*^−/−^ single mutants^20^, suggesting that NIPBL may exert deleterious transcriptional activity in the absence of its binding partner. These findings imply that MAU2 not only facilitates cohesin loading but also modulates transcriptional regulatory functions. However, the developmental consequences of *MAU2* constitutional haploinsufficiency remained so far poorly understood. Although in a previous study *Mau2*^+/−^ adults were indistinguishable from wild-type littermates^20^, our heterozygous *Mau2* knockout mouse model recapitulates core human features, including short stature and microcephaly, thereby providing direct evidence that *MAU2* haploinsufficiency disrupts mammalian growth and brain development. The discrepancy with the previous report may reflect differences in allele design and genetic background, or the lack of standardized histopathological procedures, an issue we have previously shown to be a common cause of false-negative findings in the literature^18^.

Altogether, our findings provide compelling evidence that *MAU2* should be considered a new CdLS-associated gene, while also highlighting that *MAU2* dysfunction can manifest as a distinct chromatinopathy with variable expressivity. Within our cohort, 11 out of 18 patients were confirmed as having CdLS through molecular analyses. In three patients (Patients 6, 11, and 13), the *MAU2* variants appeared to produce only milder effects, as indicated by the presence of a *MAU2*-specific episignature alone; consistent with this, none of these three patients exhibited classic CdLS features. These observations indicate that *MAU2* variants can give rise to a continuum of phenotypes, ranging from a mild *MAU2*-related syndrome to cases with partial or full overlap with CdLS, with corresponding differences in clinical presentation and methylation profiles. Together, these data support the concept of a *MAU2*-related chromatinopathy characterized by variable expressivity, encompassing both a distinct syndrome and phenotypes overlapping with CdLS.

Importantly, two variants tested negative across all molecular assays (p.(Cys394Tyr), and p.(Leu528Phe)). In line with this, the corresponding individuals exhibited low CdLS clinical scores (4 points for both patients). These findings raise two possible interpretations: either these two variants are benign and unrelated to the patients’ clinical manifestations, or they may contribute to disease through a distinct pathogenic mechanism that leads to an alternative phenotypic outcome. The shared structural eye anomalies in Patients 3 and 5—features uncommon in CdLS—support the possibility of a distinct phenotypic outcome. Beyond phenotypic overlap, several lines of evidence support the pathogenicity of these variants, including their *de novo* occurrence, strong *in silico* predictions, and classification as likely pathogenic according to ACMG criteria. These findings suggest that these two variants might be associated with an alternative pathogenic mechanism. We hypothesize that this additional pathogenic mechanism may result from the disruption of MAU2’s role in transcriptional regulation. Our previous work has shown that MAU2 is not merely a chaperone for NIPBL but also plays an active role in regulating gene expression^12^, possibly by facilitating interactions with other transcription factors. It is therefore plausible that the two non-CdLS-associated variants in our cohort impair MAU2’s ability to interact with a specific transcriptional partner. Investigating this potential mechanism could not only broaden our understanding of MAU2’s involvement in disease beyond classical CdLS, but also provide valuable insight into its poorly characterized role in transcriptional regulation.

## Materials and Methods

### MAU2 patient cohort

Our cohort was collected with the assistance of GeneMatcher^33^ and international collaborations. Exome sequencing, genome sequencing, or gene panel testing was conducted at the respective institutions. All procedures adhered to the ethical guidelines of the relevant institutional and/or national research committees, in accordance with the Helsinki Declaration and its subsequent amendments. Patient data were anonymized, and detailed clinical descriptions were provided by the referring physicians. Informed consent for participation in the research study was obtained from all individuals or their legal guardians, under locally approved IRB protocols. Additional consent was secured for the publication of photographs.

### *MAU2* variants and *in silico* predictions

Variants were mapped on the *MAU2* NM_015329.4 RefSeq transcript following HGVS recommendations^34^ and classified according to ACMG Guidelines^35^.

The pathogenicity of missense variants was assessed using a combination of multiple *in silico* prediction algorithms. The Combined Annotation-Dependent Depletion (CADD) score^36^ was calculated for all variants based on GRCh37-v1.7 genomic coordinates. Evolutionary conservation of the affected MAU2 amino acids was analyzed using Alamut Visual Plus™ (SOPHiA GENETICS, Lausanne, Switzerland). Additional prediction tools used included PolyPhen-2 (v2.2.3), REVEL (v2021-05-03), SIFT (v6.2.0), MutationTaster (v2021), and AlphaMissense^37^.

### *MAU2* Episignature Discovery

Genome-wide DNA methylation profiling was performed using the Illumina Infinium Methylation EPIC Bead Chip, following the manufacturer’s instructions (Illumina, San Diego, CA, USA). The procedures for DNA methylation analysis and episignature discovery adhered to previously established protocols^17, 38^. Intensity data were processed in R (version 4.4.2) using the sesame package for normalization and background correction^39^. Probes with detection p-value >0.01, located on sex chromosomes, near CpG interrogation sites or single nucleotide extension sites, containing single nucleotide variations, or showing cross-reactivity with other chromosomal locations other than their target regions were removed, as were arrays with >5% probe failure or those exhibiting batch effect. Genome-wide methylation density was assessed to confirm bimodal distribution, with nonconforming samples removed. Principal Component Analysis (PCA) was then used to evaluate batch structure and identify outliers.

A control cohort was randomly selected from the EpiSign Knowledge Database (EKD) and matched to *MAU2* cases according to age, sex, and array type using the MatchIt package (version 4.7.0). To establish an optimal sample size, several rounds of matching were conducted. After each iteration, PCA was performed on the case and matched control samples, and any outlier controls were subsequently removed. This process was repeated until no further outliers were detected within the first two components of the PCA.

Methylation at each probe was expressed as a β-value (0–1), computed by dividing the methylated signal intensity by the sum of methylated and unmethylated signal intensities. β-values were then logit-transformed into M-values (log2(β/(1−β))) to ensure homoscedasticity for downstream linear modeling. Differentially methylated probes (DMPs) were identified using multivariate linear regression in the limma package (version 3.62.2), adjusting for estimated blood cell composition^40^. Resulting p-values were moderated with eBayes and corrected for multiple testing using the Benjamini–Hochberg (BH) method. Probe selection was performed in three stages. First, each probe’s absolute methylation difference between cases and controls was multiplied by the negative logarithm of its adjusted p-value, and the top-ranking probes were retained. Next, from this set, the probes with the highest area under the curve (AUC) values were selected based on receiver operating curve (ROC) characteristic analysis. Lastly, probes showing pairwise Pearson correlation coefficients greater than 0.7 between cases and controls were removed to improve genome-wide representation, resulting in the final probe set. These probes were then analyzed by hierarchical clustering using Ward’s method on Euclidean distances and multidimensional scaling (MDS) to evaluate separation between cases and controls.

A binary support vector machine (SVM) classification model was developed using the e1071 R package (version 1.7.16), according to previously described procedures^17, 38^. The classifier generates methylation variant pathogenicity (MVP) scores (0–1), which provide a measure of prediction confidence for each disorder: scores close to 1 indicate a high probability that the sample’s methylation pattern corresponds to the target syndrome, while scores near 0 indicate a methylation pattern characteristic of controls. The model was trained using MAU2 cases with their matched controls, as well as 75% of other controls and 75% of samples from other rare disorders obtained from the EKD. The model was tested using the remaining 25% of control samples together with the remaining 25% of samples from other rare disorders within the EKD. Specificity was evaluated using the proportion of samples in the “Unresolved” column and an MVP threshold of 0.25.

Robustness was assessed by multiple rounds of leave-one-out cross-validation (LOOCV), where in each round one MAU2 sample was withheld for testing while the remaining MAU2 samples guided probe selection and model construction. The resulting model was then applied to classify the withheld sample. Results were visualized using heatmaps, MDS plots, and MVP plots to illustrate sample classification and the distinction between the test sample and the other samples in each round.

To evaluate whether MAU2 samples align with the known CdLS episignature, DNA methylation data were analyzed using the clinically validated EpiSign classifier, following previously established methods^17, 26, 27, 28^. Normalized and background-corrected β-values derived from the EPIC V2 arrays were processed through the established SVM model, which references the EKD of disorder-specific and control profiles to generate MVP scores (0-1). MVP scores near 1 indicate a high probability that the sample matches the CdLS signature. A positive classification is typically defined by an MVP score greater than 0.5. The final matched EpiSign result is generated using these scores, along with the assessment of hierarchical clustering and multidimensional scaling.

### Mammalian two-hybrid assay

A fragment of NIPBL containing amino acids 1–300 was inserted into the pCMV-BD expression plasmid (#211342, Agilent Technologies, Santa Clara, CA, USA). The full-length open reading frame of MAU2 was cloned into the pCMV-AD plasmid (#211343, Agilent Technologies). MAU2 mutant constructs were generated through site-directed mutagenesis based on divergent PCR in combination with 5’-phosphorylated primers. Constructs harboring the following variants were generated for this assay: p.(Cys50Ser), p.(Val54_Pro56del), p.(Ala309_Lys322del), p.(Gln310_Ala316del), p.(Lys322del), p.(Ala378_Gln380delinsAsp), p.(Leu381Pro), p.(Cys394Tyr), p.(Leu528Phe), p.(Asp534His), as well as the synonymous variant, frequent in the healthy population, p.(Thr355Thr). HEK293 cells were transiently transfected in 24-well plates with FuGene-HD transfection reagent (E2311, Promega, Madison, WI, USA), according to the manufacturer’s instructions. Each well was transfected with 250 ng of the pCMV-BD-NIPBL 1-300aa, 250 ng of the pCMV-AD-MAU2 wild type or mutant constructs, 250 ng of the Firefly Luciferase reporter plasmid and 2,5 ng of the phRG-TK Renilla luciferase expression plasmid. Activity of Firefly and Renilla luciferases was measured 24 hours post-transfection with the Dual Luciferase Reporter Assay System (E1980, Promega) using the GloMax Discover System (Promega). All measurements were performed in triplicate in at least six independent experiments. Relative luciferase activity, indicating the strength of the interaction, was determined as the triplicate average of the ratio between the Firefly and the Renilla luciferase activity.

The statistical significance of the mammalian two-hybrid results was first assessed using the Kruskal-Wallis test to evaluate differences across groups. After identification of a significant effect, pairwise comparisons were performed using Dunn’s test, with a Bonferroni correction applied to account for multiple comparisons.

### Expression controls of mammalian two-hybrid constructs

Expression of exogenous MAU2 wild type and mutant constructs (pCMV-AD-MAU2 and the in-house generated 3xFLAG-MAU2) was verified by Western Blot. HEK293 cells were transiently transfected in 10 cm plates with the jetOPTIMUS® DNA Transfection Reagent (101000006, Polyplus, Illkirch, France), following the manufacturer’s instructions. Each plate was transfected with 10 µg of the corresponding MAU2 construct. Cells were harvested 24 hours after transfection. After resuspension in 250 µl of lysis buffer (150 mM NaCl, 20 mM NaF, 50 mM Tris pH 7.5, 0.1% NP40, 1 mM EGTA, protease inhibitor cocktail (04693132001, Merck, Darmstadt, Germany)), samples were incubated on ice for 60 minutes. Protein lysates were obtained by high-speed centrifugation at 4°C for 30 minutes. Protein concentration was determined using the BCA Protein Assay Kit (#23225, Pierce, Rockford, IL, USA) following the manufacturer’s protocol. Protein expression was analyzed by Western blotting using Mini-PROTEAN® TGX™ gels (4568106, Bio-Rad, Hercules, CA, USA). Briefly, 20 µg of each protein lysate were separated by SDS-PAGE in 1X Tris-Glycine-SDS running buffer and transferred to a nitrocellulose membrane using a semi-dry transfer system (Trans-Blot® SD Semi-Dry Transfer Cell, Bio-Rad) at 25 V for 30 minutes, following the manufacturer’s protocol. After transfer, the membrane was blocked with EveryBlot Blocking Buffer (#12010020, Bio-Rad) for 15 minutes at room temperature.

Membranes were subsequently incubated overnight at 4°C with the following primary antibodies: MAU2 (1:1000; ab183033, Abcam, Cambridge, MA, USA), NF-kB (1:500; sc-372, Santa Cruz Biotechnology, Dallas, TX, USA), FLAG (1:1000; F3165, Sigma-Aldrich, St. Louis, MO, USA), and GAPDH (1:1000, 2118S, Cell Signaling Technology, Danvers, MA, USA). Antibodies were diluted in 5% bovine serum albumin (BSA) in Tris-buffered saline with 0.1% Tween-20 (TBST). Following incubation, the membrane was washed four times with TBST, each wash lasting ten minutes. It was then incubated for 1 hour at room temperature with the appropriate HRP-conjugated secondary antibody, diluted in the same buffer used for the primary antibodies: goat-α-rabbit (#31460, Thermo Fisher Scientific, Waltham, MA, USA), diluted 1:20,000 for the MAU2- and NF-kB membranes and 1:200,000 for the GAPDH-membrane, and goat-α-mouse (#31430, Thermo Fisher Scientific), diluted 1:100,000 for the FLAG-membrane. Following four washes with TBST and two washes with TBS, protein bands were detected using an enhanced chemiluminescence (ECL) substrate (SuperSignal West Femto ECL, #34095, Thermo Fisher Scientific) and visualized with the Intas ChemoStar Touch imaging system (Intas Science Imaging Instruments, Göttingen, Germany).

### RNA isolation, cDNA synthesis and Real-Time PCR

RNA extraction was carried out with the ReliaPrep™ RNA Cell Miniprep System (Promega) according to the manufacturer’s instructions. The LunaScript® RT SuperMix Kit (E3010L, New England Biolabs, Ipswich, MA, USA) was used to retro-transcribe 1 µg of RNA. cDNA synthesis was performed in two independent reactions for each of the three biological replicates of the same sample.

The expression levels of the transcripts of interest were assessed using the qPCRBIO Probe Mix Hi-ROX assay for Real-Time PCR (PB20.23-05, PCR Biosystems, London, UK). The investigation was run on the LightCycler 480 (Roche, Basel, Switzerland). The following TaqMan gene expression assays were used for the analysis: Hs01062386_m1 (for the *MAU2* transcript) and Hs00209846_m1 (for the *NIPBL* transcript) (Thermo Fisher Scientific). The *GAPDH* and *NADH* genes were selected as endogenous normalizer and amplified with the TaqMan gene expression assay ID Hs02758991_g1 and hs00190020_m1 (Thermo Fisher Scientific). Relative gene expression was determined using the ΔΔCt method^41^. Mann-Whitney U tests, followed by Bonferroni correction for multiple comparisons, were used to assess the statistical significance of the qPCR results. Pairwise comparisons were made between each experimental group and the control group.

### Western Blot on patients’ fibroblasts

Fibroblast pellets were resuspended in 100 µl of RIPA lysis buffer (#89900, Thermo Fisher Scientific) supplemented with protease inhibitor cocktail (04693132001, Merck). Samples were incubated on ice for 30 minutes. Protein lysates were then obtained by high-speed centrifugation at 4°C for 30 minutes. Western Blot experiments were carried out as previously described in the “Expression Controls for mammalian two-hybrid constructs” section unless otherwise specified. The following primary antibodies were used for detection: MAU2 (1:1000; ab183033, Abcam), Actinin (1:1000; sc-17829, Santa Cruz Biotechnology), and NIPBL (1:500; KT54, Absea). The secondary antibodies were diluted as follows: goat-α-rabbit for MAU2 (1:10,000; #31460, Thermo Fisher Scientific), goat-α-mouse for Actinin (1:100,000; #31430, Thermo Fisher Scientific), and goat-α-rat for NIPBL (1:10,000; #A10549, Thermo Fisher Scientific).

### Generation and characterization of the *Mau2^tm1b(KOMP)Wtsi^* knockout mouse

The *Mau2^tm1b(KOMP)Wtsi^* mouse was generated at the Wellcome Sanger Institute on a pure C57BL/6N genetic background by homologous recombination in embryonic stem cells using the knockout-first allele method^19^. In this strategy, an exon common to all transcripts (exon 5) was targeted, upstream of which a LacZ cassette was inserted. Exon 5 of the *Mau2* allele, flanked bilaterally by *loxP* sequences, was deleted using Cre recombinase that recognizes *loxP* sites, thereby producing the *Mau2^tm1b(KOMP)Wtsi^*knockout allele.

Through collaboration with the Sanger Institute Mouse Genetics Project, we obtained weight and body height measurements of seven *Mau2^+/−^* female mice and 340 baseline WT female mice, as well as seven *Mau2^+/−^* male mice and 345 baseline WT male mice. We also received brain samples from *Mau2^+/−^* mice and performed comprehensive neuroanatomical studies on three *Mau2^+/−^* mutants and 44 baseline WT mice at 16 weeks of age, as previously described^21, 22^. A validated statistical model (G*Power) was applied to detect neuroanatomical defects with an effect size ≥10% at 80% power^18^. Brains were fixed in 4% buffered formalin for 48 hours, transferred to 70% ethanol, and paraffin-embedded. Coronal sections (5 μm thickness) were obtained at Bregma +0.98 mm and Bregma –1.34 mm (Allen Mouse Brain Atlas) using a Leica RM 2145 microtome. Sections were double-stained with 0.1% Luxol Fast Blue (Solvent Blue 38; Sigma-Aldrich) and 0.1% Cresyl violet acetate (Sigma-Aldrich), then scanned at 20× resolution using the Nanozoomer whole-slide scanner 2.0HT C9600 series (Hamamatsu Photonics, Japan). Images were quality controlled for sectioning accuracy, asymmetries, and histological artifacts. All samples were assessed for cellular ectopia. 63 brain parameters (comprising left and right hemispheres) were measured blind to the genotype across two coronal sections as described in Mikhaleva *et. al.*^21^. Data were analyzed using a t-test to determine whether a brain region was associated with neuroanatomical defect or not.

### Use of AI Tools in Data Analysis and Manuscript Preparation

ChatGPT (OpenAI, San Francisco, CA, USA) was used to assist in data analysis (R scripting) and to provide language editing suggestions for this manuscript. The authors reviewed the output for accuracy.

## Data Availability

All data produced in the present work are contained in the manuscript

## Declarations

## Ethics approval and consent to participate

Informed consent for participation in the research study and permission for data publication was obtained from all individuals or their legal guardians, under locally approved IRB protocols. The project was approved by the Ethic Committee of the University Hospital Essen (UME-ID-12571). Additional consent was secured for the publication of photographs.

## Availability of data and material

All variants have been submitted to the ClinVar Database. The submission number is SUB15713355. Accession numbers will be provided prior to publication.

## Authors’ contributions

Conceptualization: I.P., F.J.K. Investigation: I.P., A.H., M.G.S., S.C.C., L.D., D.K., J.B., J.K., L.S., J.E., R.K., B.Y., and K.S.W. Formal analysis: I.P., A.H., M.G.S., L.D., S.C.C., J.K., E.L., B.S., B.Y., and K.S.W. Resources: J.W., P.M.B, E.M.K., L.A., H.A., F.S.A., S.E.A., D.B., K.C., L.C., A.R.C., A.A.L.J., A.C.L., B.D., P.J., A.O.K., K.K., G.J.L., E.M., J.M.C., L.K.K., S.M., G.N., Y.N., T.O., J.Pi., J.Pr., B.P., F.J.R., E.Ra., C.R., E.Ru., S.Sa., H.E.S., S.St., E.A.S., and A.K. Visualization: I.P., A.H., M.G.S., and L.D. Writing – original draft: I.P., A.H., B.Y., K.S.W., and F.J.K. Writing – review and editing: All authors reviewed and edited the manuscript.

## Acknowledgements

We are deeply grateful to the patients and their families for their trust, willingness to share clinical data, and generous contribution of time and samples, without which this study would not have been possible.

This study made use of data generated by the International Mouse Phenotyping Consortium (IMPC, https://www.mousephenotype.org). We gratefully acknowledge the IMPC and its member institutions for their efforts in generating and providing access to phenotyping data. In particular, we are extremely grateful to Valérie E. Vancollie and Christopher J. Leliott, as well as members of the Sanger Institute Mouse Pipelines teams and the Research Support Facility, for the production and management of the *Mau2^+/−^* mouse line used in this study. We thank Dorinda Wright, Susan Mary Smith, and Lindsey Proctor for histological work at HistologiX Ltd, and the students and technicians of the Yalcin’s laboratory for their contributions to mouse phenotyping, in particular Kevin Navarro, Helen Whitley, Christel Wagner, Marie-Christine Fischer, Anna Mikhaleva, Rebecca Balz, and Léo Gagliardi. A.K., F.J.K. (Essen, Germany), and B.Y. (Dijon, France) are members of the European Reference Network (ERN) ITHACA. P.M.B. was supported by award K08NS117891 from the U.S. National Institute of Neurological Disorders and Stroke (NINDS) and an award from the Boston Children’s Hospital Office of Faculty Development.

