## Supplementary Figures for "Pathogenic variants in the cohesin loader subunit MAU2 lead to a new Cornelia de Lange Syndrome subtype"

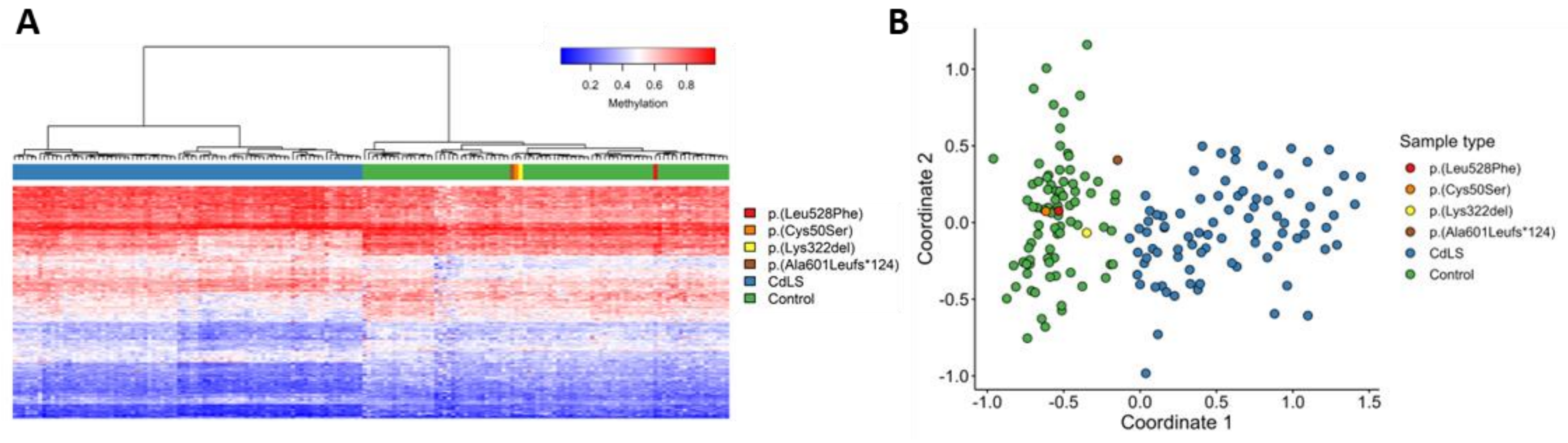


**Supplementary Figure 1. EpiSign (DNA methylation) analysis of *MAU2* cases negative for the CdLS episignature.** (A) Hierarchical clustering demonstrates that the four cases negative for the CdLS episignature (red, orange, yellow, brown) exhibit a DNA methylation profile similar to controls (green), and distinct from subjects with a confirmed CdLS episignature when plotted alone (blue). (B) Multidimensional scaling analysis reveals that the four cases negative for the CdLS episignature (red, orange, yellow, brown) cluster with controls (green) and are distinct from the confirmed CdLS cohort (blue).

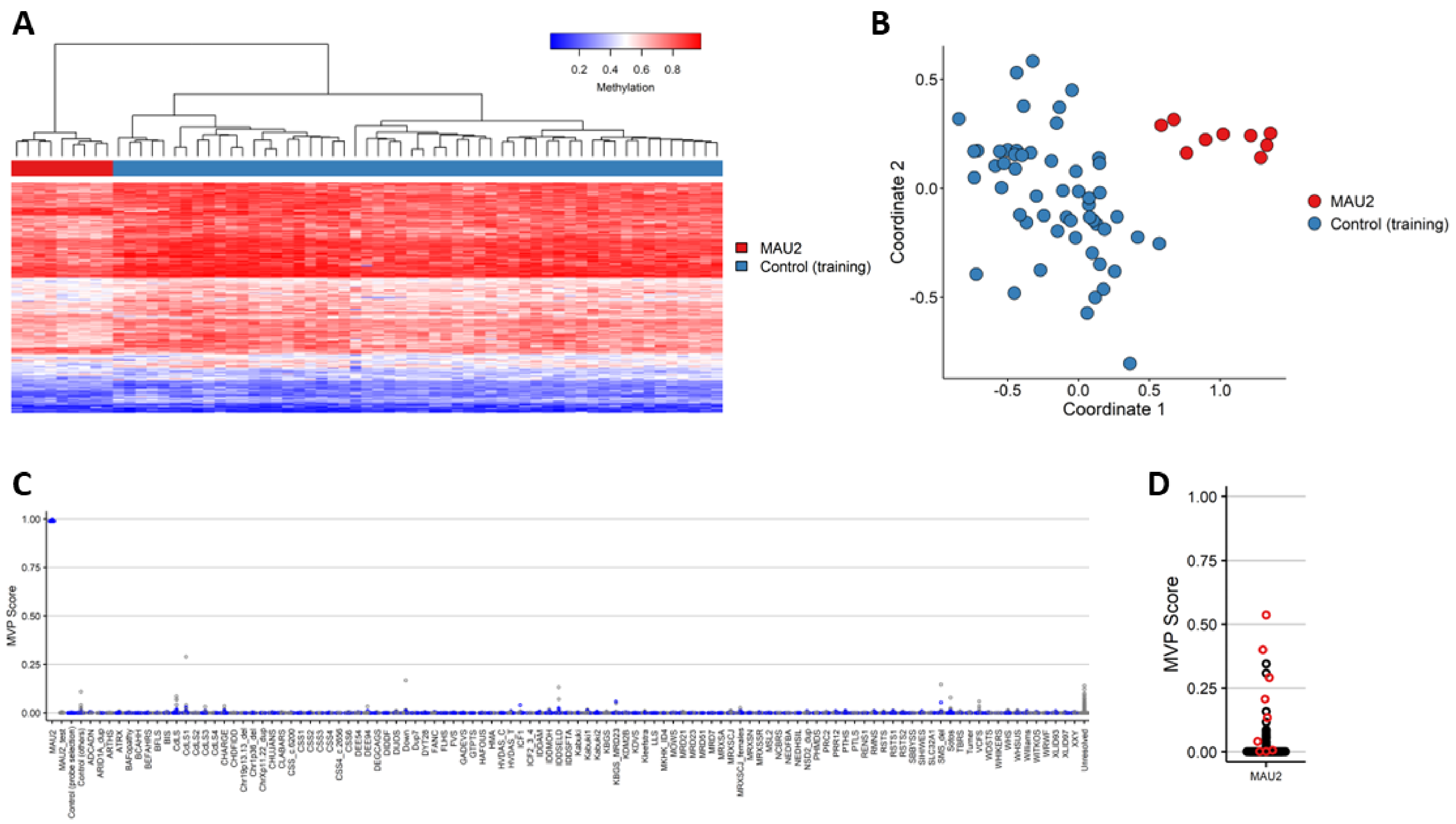


**Supplementary Figure 2. Identification and evaluation of the *MAU2* episignature using nine samples.** (A) Euclidean hierarchical clustering (heatmap): each column represents a single *MAU2* case or a matched control, and each row corresponds to one of the 211 CpG probes selected for episignature discovery. *MAU2* cases (red) and controls (blue) cluster separately, with *MAU2* cases exhibiting a methylation pattern distinct from controls. (B) Multidimensional scaling (MDS) plot showing separation between *MAU2* cases (red) and controls (blue). (C) MVP scores calculated by the SVM classifier are plotted, with training samples shown as blue circles and testing samples as gray circles. The model was trained using the *MAU2* cases, their matched controls, and 75% of additional controls and 75% of other disorder samples from the EKD, while the remaining 25% of controls and 25% of disorder samples were used as the testing set. MVP scores for *MAU2* cases are near 1, while the remaining training and testing samples cluster near 0, indicating specificity for the classifier. (D) Summary MVP plot of nine rounds of leave-one-out cross validation (LOOCV) of the *MAU2* classifier. For each round of LOOCV, an MVP score was generated for the withheld sample. The MVP scores for the *MAU2* samples (red) were then plotted alongside the average MVP scores across all LOOCV rounds for other samples in the EKD (black), including controls and individuals with different episignature disorders.

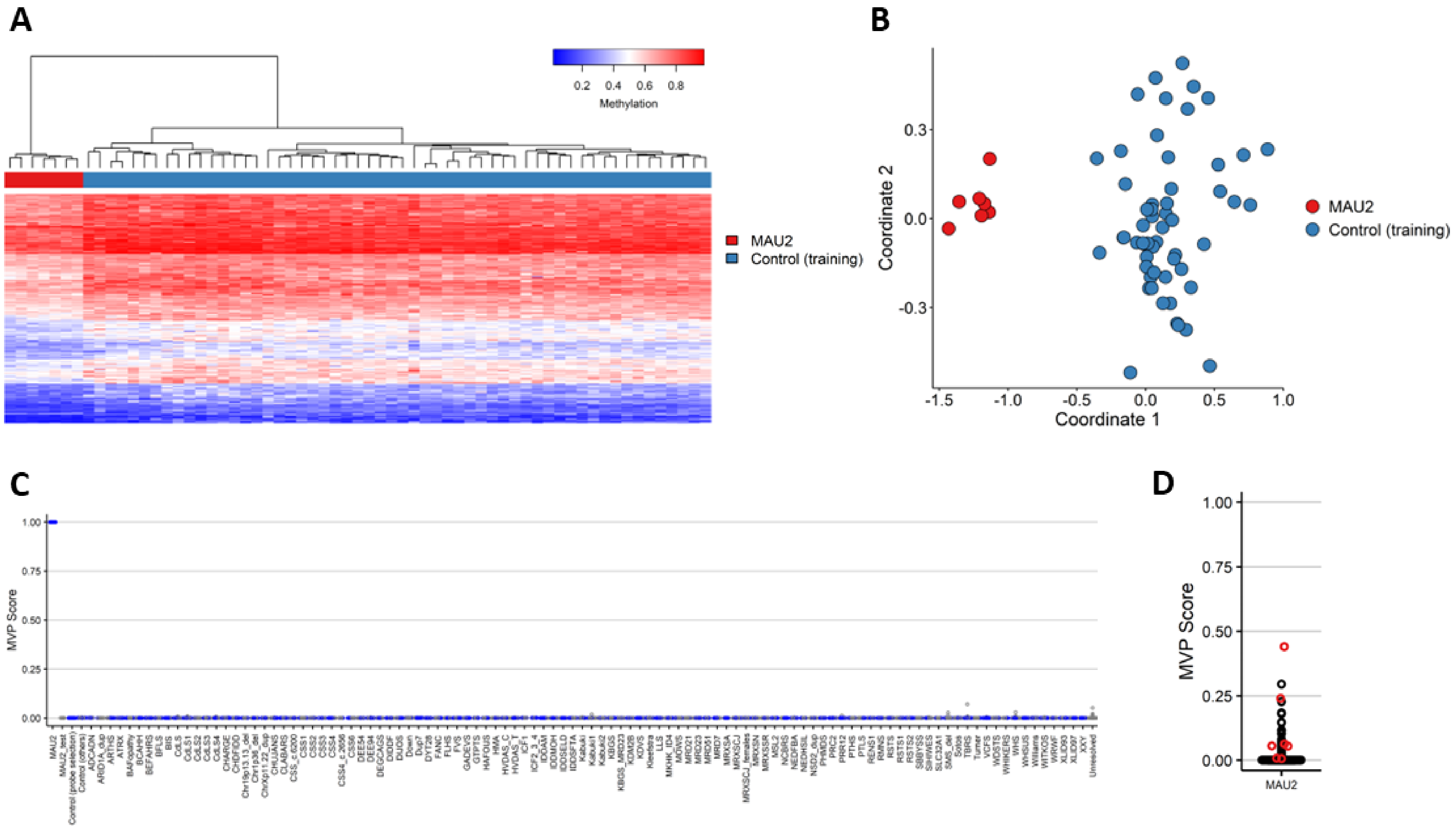


**Supplementary Figure 3. Identification and evaluation of the *MAU2* episignature using seven samples.** (A) Euclidean hierarchical clustering (heatmap): each column represents a single *MAU2* case or a matched control, and each row corresponds to one of the 203 CpG probes selected for episignature discovery. *MAU2* cases (red) and controls (blue) cluster separately, with *MAU2* cases exhibiting a methylation pattern distinct from controls. (B) Multidimensional scaling (MDS) plot showing separation between *MAU2* cases (red) and controls (blue). (C) MVP scores calculated by the SVM classifier are plotted, with training samples shown as blue circles and testing samples as gray circles. The model was trained using the *MAU2* cases, their matched controls, and 75% of additional controls and 75% of other disorder samples from the EKD, while the remaining 25% of controls and 25% of disorder samples were used as the testing set. MVP scores for *MAU2* cases are near 1, while the remaining training and testing samples cluster near 0, indicating specificity for the classifier. (D) Summary MVP plot of seven rounds of leave-one-out cross validation (LOOCV) of the *MAU2* classifier. For each round of LOOCV, an MVP score was generated for the withheld sample. The MVP scores for the *MAU2* samples (red) were then plotted alongside the average MVP scores across all LOOCV rounds for other samples in the EKD (black), including controls and individuals with different episignature disorders.

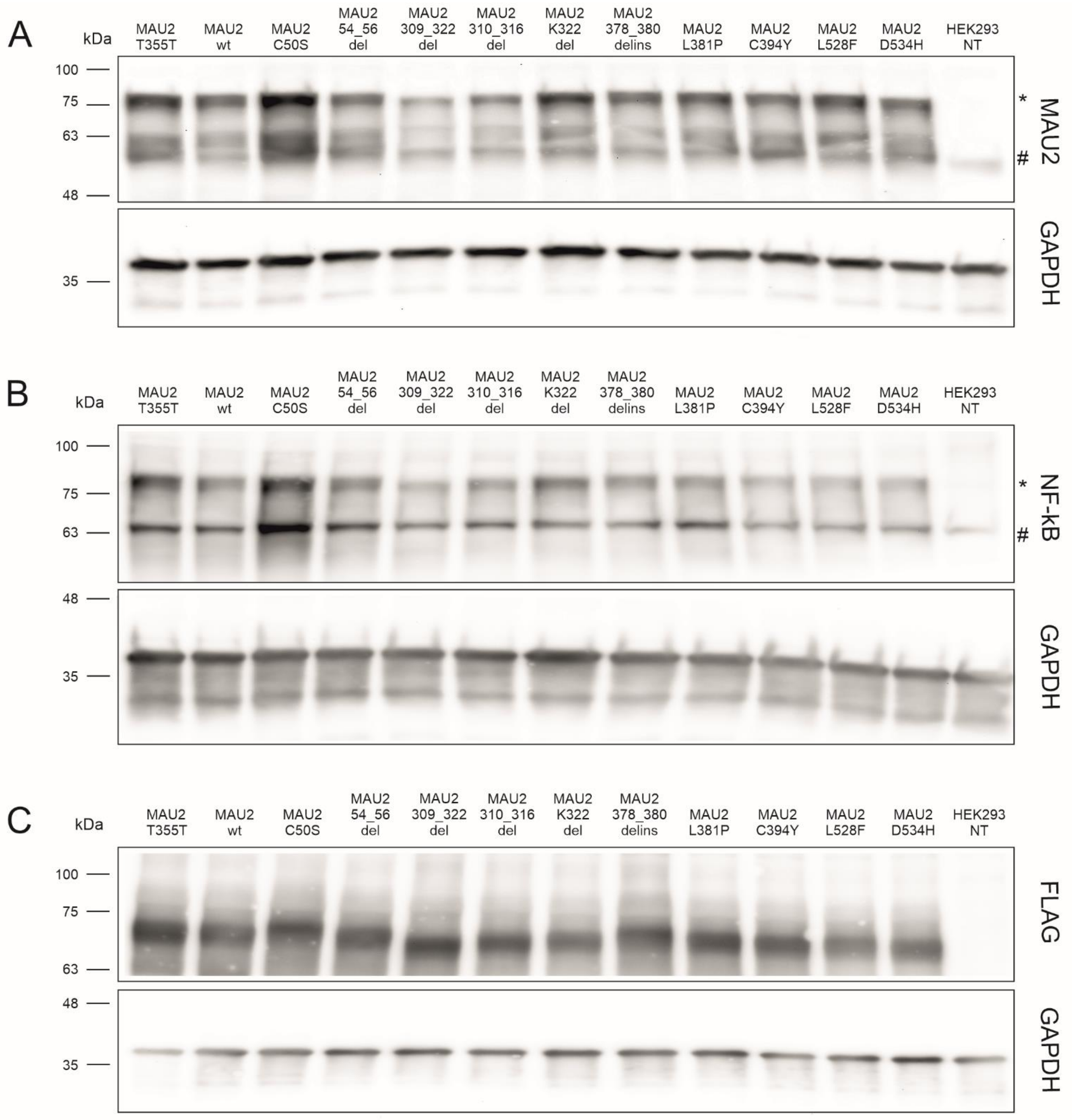


**Supplementary Figure 4. Expression validation of MAU2 constructs (refers to Figure 3).** (A) Confirmation of expression for pCMV-AD_MAU2 constructs used in the mammalian two-hybrid assay. Constructs were detected with an anti-MAU2 antibody, which recognizes both endogenous MAU2 (#) and the exogenously tagged MAU2 (*).
(B) Confirmation of expression for pCMV-AD_MAU2 constructs used in the mammalian two-hybrid assay. Constructs were detected with an anti-NF-κB antibody, which recognizes both endogenous NF-κB (#) and the exogenous NF-κB-MAU2 fusion construct (*).
(C) Confirmation of expression for 3xFLAG-MAU2 constructs. Constructs were detected using an anti-FLAG antibody.
In all panels, GAPDH was used as a loading control.

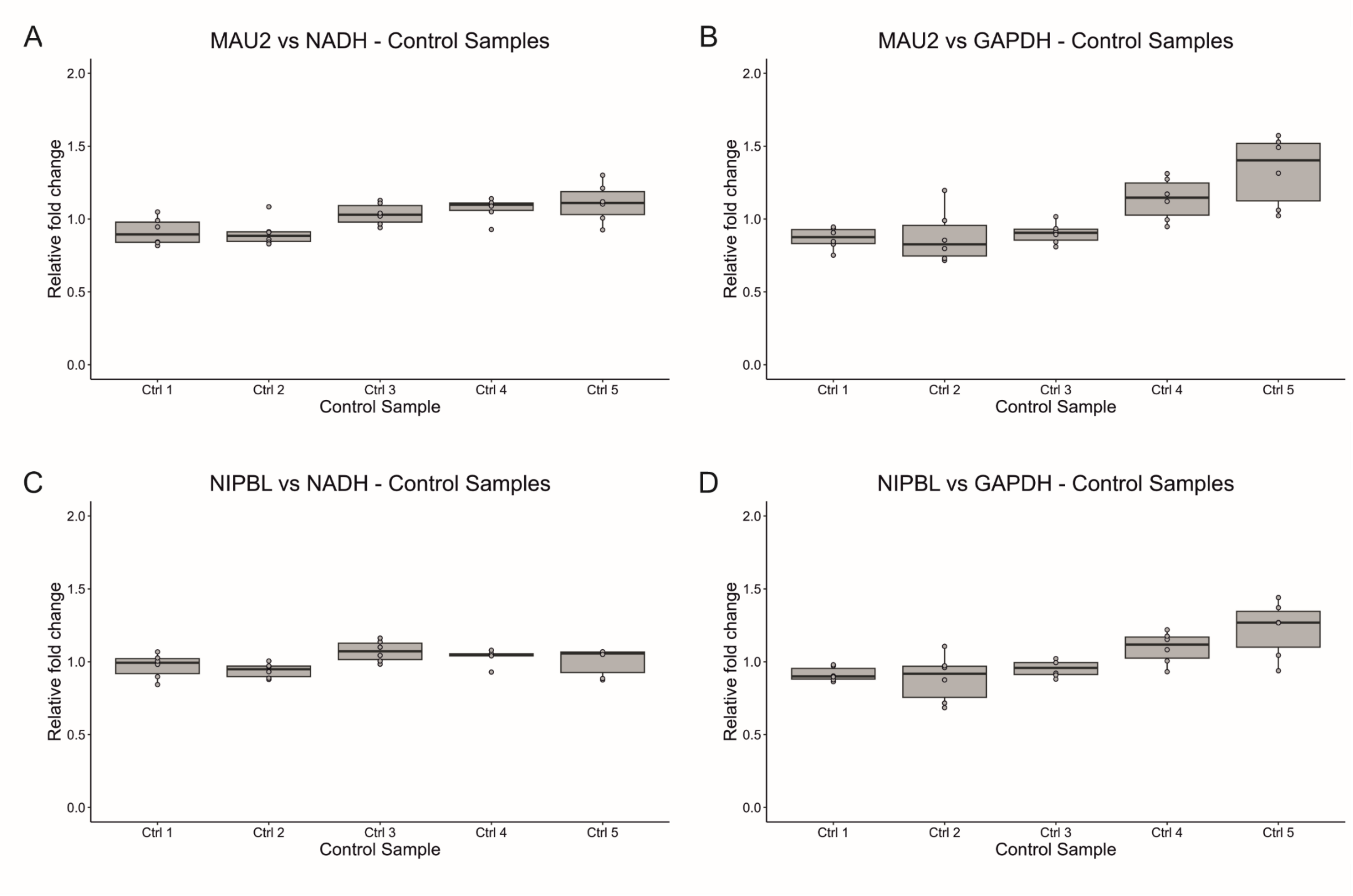


**Supplementary Figure 5. Distribution of *MAU2* and *NIPBL* expression across controls (refers to Figure 4).** (A) *MAU2* expression levels relative to *NADH* in five healthy unrelated controls used to normalize the expression levels in patients. (B) *MAU2* expression levels relative to *GAPDH* in five healthy unrelated controls used to normalize the expression levels in patients. (C) *NIPBL* expression levels relative to *NADH* in five healthy unrelated controls used to normalize the expression levels in patients. (D) *NIPBL* expression levels relative to *GAPDH* in five healthy unrelated controls used to normalize the expression levels in patients.

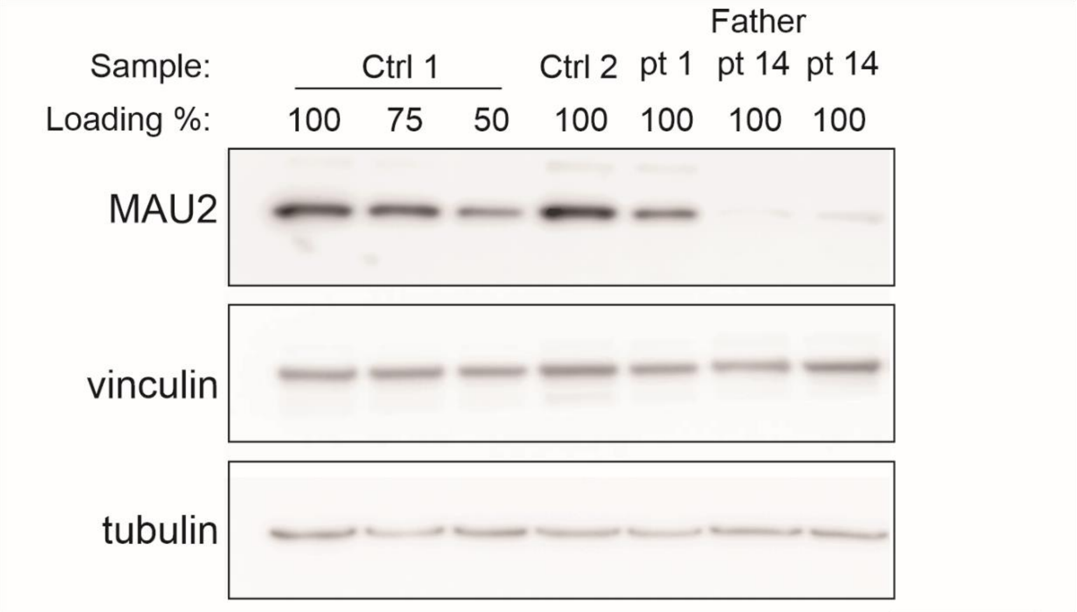


**Supplementary Figure 6. Fibroblasts with a frameshift variant exhibit MAU2 haploinsufficiency (refers to Figure 4)**. Three different amounts of control 1 protein were loaded to confirm the linearity of detection. Reduced MAU2 protein levels are evident in patient 14 and her father, consistent with haploinsufficiency, but not in patient 1. Vinculin and tubulin served as loading controls. MAU2 was detected using a rabbit anti-MAU2 antibody (Abcam, ab46906).
